# Sleep as a window into thalamocortical pathology: generative modeling implicates NMDA receptor hypofunction in 22q11.2 deletion syndrome

**DOI:** 10.1101/2025.08.01.25332824

**Authors:** Lioba C S Berndt, Rosina M Diebel, Nicholas A Donnelly, Jeremy Hall, Marianne B M van den Bree, Rick A Adams, Alexander D Shaw, Matt W Jones

## Abstract

22q11.2 deletion syndrome (22q11.2DS) is a strong genetic risk factor for neuropsychiatric conditions, including schizophrenia, yet the underlying synaptic mechanisms remain unclear. Sleep EEG suggests thalamocortical dysfunction, but scalp data alone lack mechanistic resolution. Computational modelling can bridge this gap by inferring receptor-level dynamics from EEG.

We applied a conductance-based thalamocortical Dynamic Causal Model (DCM) to sleep–wake EEG from children with 22q11.2DS (*n* = 28) and their neurotypical siblings (*n* = 17), estimating contributions of AMPA, NMDA, GABA_A_, and GABA_B_ conductances. Building on these estimates, we investigated which receptor systems, if perturbed, could shift circuit dynamics toward sibling patterns. To address this, we implemented *in silico* pharmacology by systematically scaling receptor-mediated conductances. Increasing NMDA receptor (NMDA-R) efficacy consistently produced the strongest improvements in alignment with sibling spectra (effect size = 0.32 in light NREM, 0.45 in deep NREM), whereas AMPA- or GABA-based manipulations were weaker. The most influential pathways were recurrent NMDA-R excitation among superficial pyramidal cells and NMDA-R excitation from spiny stellates to superficial pyramidal populations. Exploratory regressions linked greater thalamocortical delay during deep sleep to greater sleep problems (*β* = 0.34, *p*_*FDR*_ = 0.006), and AMPA-mediated excitation of interneurons during wakefulness to anxiety symptoms (*β* = −0.32, *p*_*FDR*_ = 0.032).

These findings implicate NMDA receptor hypofunction as a key mechanism in 22q11.2DS and suggest it may serve as a treatment target. We further show that DCM-based virtual pharmacology can simulate drug-level interventions, and we are now testing whether NMDA receptor modulation restores network activity in preclinical models (e.g., mouse models of 22q11.2DS).

## 1 Introduction

22q11.2 deletion syndrome (22q11.2DS) is a recurrent microdeletion syndrome with an estimated prevalence of 1:3000 live births. It confers substantial vulnerability to intellectual disability, autism spectrum disorder (ASD), attention-deficit hyperactivity disorder (ADHD), and epilepsy^1,2,3^. Notably, 22q11.2DS represents the strongest known genetic risk factor for schizophrenia, with up to 25% of adults getting diagnosed with schizophrenia and 40% developing psychotic conditions more broadly^4,5,6^. Children and young people with this deletion afford a unique opportunity to examine psychosis risk before onset, prior to the confounding effects of disease progression and antipsychotic medication on phenotypic manifestations and their comorbidities.

Recent research has highlighted disrupted sleep physiology as a key factor in the psychiatric risk associated with 22q11.2DS^7^. Sleep disturbances, including insomnia and sleep fragmentation, are prevalent in 22q11.2DS and co-occur with distinct neural signatures during sleep. For example, altered spindle-slow-wave coupling correlates with the severity of psychopathology^8^. These neurophysiological signatures may reflect cross-diagnostic patterns observed in schizophrenia, ASD, and ADHD^9,10,11,12,13,14^, disorders that may share some neurobiological similarities with 22q11.2DS. Such parallels suggest shared mechanisms in sleep-related circuit dysfunction that may transcend current diagnostic boundaries.

To systematically investigate these mechanisms, we established the Sleep Detectives initiative, a multidisciplinary program examining sleep (behaviour and neurophysiology) and cognition in 22q11.2DS and other Copy Number Variants associated with increased risk of psychosis. Our work builds on the findings from a prior sleep EEG study^8^ which demonstrated distinct neural alterations during sleep in individuals with 22q11.2DS compared to sibling controls, including elevated slow-wave and spindle amplitude, as well as stronger spindle-slow-wave coupling. These EEG patterns partially mediated the impact of the deletion on anxiety, ADHD, and ASD symptoms. While these results confirm fundamental differences in neural circuit function, conventional EEG analyses cannot fully capture the underlying synaptic mechanisms, which would have direct implications for both early detection and targeted treatment development, as well as for bridging the gap between genetic risk factors, neurobiological mechanisms, and phenotypic expression in neurodevelopmental disorders.

To address this gap, we turned to neural mass modelling, which provides a mechanistic framework for understanding population-level neural dynamics. These models formalise how synaptic interactions generate emergent oscillatory patterns^15^. Unlike descriptive (i.e. phenomenological) approaches, neural mass models parameterise physiological properties (e.g., synaptic strengths, time constants) that define a mechanistic landscape of circuit alterations. Dynamic Causal Modelling (DCM) operationalises this framework, inferring hidden synaptic parameters from EEG data by optimising model-generated activity to match observed spectra^16,17^. To pinpoint synaptic mechanisms underlying sleep disturbances in 22q11.2DS, we employed conductance-based DCM^16^. This approach integrates Hodgkin-Huxley-type equations^18^ to simulate thalamocortical dynamics, explicitly parameterising thalamic and cortical populations and synaptic connectivity^19^. We focused on the thalamus given its dual role in sleep regulation—orchestrating spindles and slow waves^20,21^—and its implication in schizophrenia pathophysiology, where thalamic volume reductions and disrupted connectivity are hallmarks^22,23^, making it particularly relevant for examining early markers of psychosis risk through sleep-related measures in 22q11.2DS.

Building on this generative modelling framework, we asked whether synaptic alterations in 22q11.2DS sleep neurophysiology could be targeted through receptor-specific manipulations. We used Dynamic Causal Modelling to simulate in silico “virtual pharmacology”, systematically perturbing receptor-level parameters to identify adjustments that shift 22q11.2DS neural activity toward patterns observed in neurotypical siblings. This approach offers a mechanistic step toward identifying candidate intervention targets in high-risk neurodevelopment.

## 2. Materials and methods

This study extends the findings of Donnelly et al. (2022)^8^ by applying DCM to a subset of the Cardiff University ECHO study dataset, comprising children with 22q11.2 deletion syndrome (*n=28*, age: *M=14.6, SD=3.4*) and sibling controls (*n=17*, age: *M=13.7, SD=3.4*), who took part in a study on sleep involving overnight ambulatory EEG collection^24,8^. While the original study identified distinct sleep patterns (figure 1, section 7.2 in the supplementary materials) and their psychiatric correlates such as anxiety, we now leverage these findings to explore the underlying thalamocortical dynamics through DCM. A detailed description of the original study procedures can be found in section 7.1 of the supplementary materials.

**Figure 1.**
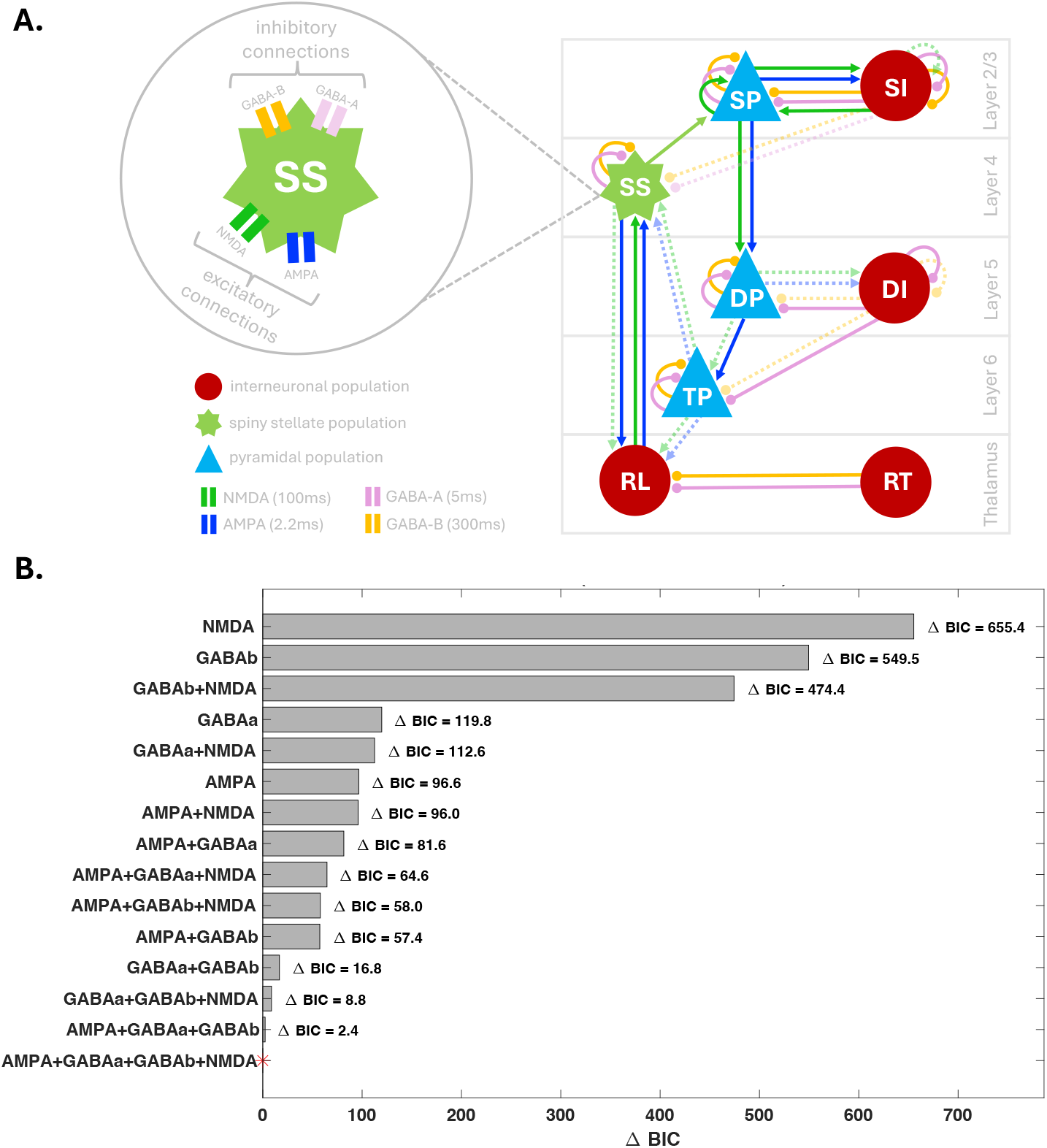
Bayesian Model Comparison. **A.** Neural circuit architecture of the winning model, illustrating the interactions between different neuronal populations through multiple receptor types. Distinct neural populations include: superficial pyramidal cells (SP) and interneurons (SI) in layer 2/3, spiny stellate cells (SS) in layer 4, deep pyramidal cells (DP) and interneurons (DI) in layer 5 and thalamic projections (TP) neurons in layer 6. These cortical populations interact with thalamic relay (RL) and reticular (RT) nuclei. Synaptic connections are mediated by four receptor types with distinct time constants: NMDA (100ms, green), AMPA (2.2ms, blue), GABA-A (5ms, pink), and GABA-B (300ms, yellow). Non-recoverable parameters were fixed to their priors (dotted + transparent). **B**. Model comparison results showing ΔBIC values for different receptor combinations in comparison to the winning model. Models are ordered by their evidence, with lower ΔBIC values indicating better model performance.

### 2.1 Computational Modeling

We employed a series of fifteen nested thalamocortical models^19^ with increasing complexity to characterise spectral activity in sleep EEG data at sensor Cz. These models systematically varied in their parameterisation of synaptic connections to identify the minimal architecture necessary for capturing the observed dynamics. We fitted the model individually for each participant (both 22q11.2DS and control siblings) to capture person-specific dynamics. This approach allowed us to make group-level inferences while accounting for individual variability.

The 15 models were constructed by systematically including or excluding connections mediated by four key synaptic receptor types (AMPA, NMDA, GABA-A, GABA-B) between eight key neuronal populations: pyramidal cells and interneurons in the superficial layers, spiny stellate cells, pyramidal cells and deep interneurons in deep layers, thalamic projection neurons, thalamic relay, and reticular nucleus. We identified the optimal model using Bayesian Information Criterion and complementary model comparison metrics that balance model fit against complexity.

To ensure interpretability of the estimated parameters, we additionally performed parameter recovery analyses, which confirmed that key synaptic parameters could be robustly estimated from the observed data. Full details of the modelling framework, model selection criteria, and parameter recovery analyses are provided in section 7.3.2 of the supplementary materials.

### 2.2 Identification of Computational Parameters in 22q11.2DS

Group differences in computational model parameters of the winning model between 22q11.2DS and siblings were identified using Parametric Empirical Bayes (PEB)^25,26^, a hierarchical Bayesian implementation of general linear models that accounts for parameter uncertainty and covariance in group-level inferences. In the supplementary materials 7.4, we detail why PEB provides more robust group-level inferences compared to frequentist statistical methods (e.g., t-tests) and, thus, is the method of choice here.

### 2.3 Mechanism Identification Through Receptor-Wide Perturbations

Having identified group-level differences in synaptic parameters, we next simulated receptor-specific synaptic perturbations to test whether biologically plausible changes could shift 22q11.2DS sleep neurophysiology toward patterns observed in neurotypical siblings. This *in silico “virtual pharmacology”* analysis focused on modulating the gain of each receptor system, mirroring the systemic action of pharmacological interventions that typically target specific receptors (but not, for example, specific neuron types or specific cortical layers).

To this end, we defined four global receptor gain parameters (c.f. hyperparameters), Γ = {*γ*_AMPA_, *γ*_NMDA_, *γ*_GABA-A_, *γ*_GABA-B_}. For each receptor *k*, we applied a uniform scaling factor *γ* ∈ [0, 2] (in steps of 0.2) to all synaptic connections mediated by that receptor:

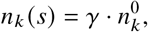

where 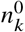 denotes the baseline conductance value, and *s* indexes simulations. All other receptor systems were held at baseline values. For each perturbation, we simulated power spectral densities across participants, computed the group-averaged PSD for the 22q11.2DS group, and quantified alignment with the sibling control PSD using mean squared error (MSE). A model effect size was computed by comparing the MSE of the perturbed model to that of the baseline model. Statistical significance was evaluated using permutation testing with 1,000 permutations^27^ (see supplemenatry section 7.5).

To determine whether the effects of receptor-wide perturbations were driven by specific synaptic pathways, we then conducted a complementary parameter-specific analysis. Let *P* = {*p*_1_, *p*_2_,…, *p*_*n*_} be the set of receptor-mediated synaptic parameters in the model (e.g., the NMDA conductance from spiny stellate to superficial pyramidal cells). For each parameter *p*_*i*_ ∈ *P*, we applied a multiplicative scaling factor *α* ∈ [0, 2] to the baseline value:

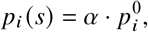

while keeping all other parameters fixed. This procedure was repeated across participants and simulations. For each perturbation, we again evaluated the group-level PSD and calculated MSE relative to controls to identify the most impactful individual connections.

### 2.4 Associations Between Thalamocortical Parameters and Psychiatric and Cognitive Measures

In addition, we examined how thalamocortical model parameters (independent variables) predict psychiatric and cognitive symptoms (dependent variables) in the 22q11.2DS group. The dependent variables comprised two categories: (1) psychiatric symptoms, including symptoms of ADHD, anxiety, ASD, sleep disturbances, and psychotic experiences (supplementary materials 7.1.2), and (2) cognitive measures, including the Cambridge Neuropsychological Test Automated Battery (CANTAB)^28^ and the Wisconsin Card Sorting Test (WCST) (details in the supplementary section 7.1.3 and 7.1.4). To manage the high dimensionality of the parameter space, we applied LASSO regression^29^, employing 5-fold cross-validation to determine the optimal penalty parameter (*λ*). This regularisation method penalises less informative predictors by shrinking their coefficients to zero, effectively isolating the most relevant parameters. For the final models, we selected the appropriate regression approach based on each measure’s distribution characteristics (supplementary materials 7.7) and included age and sex as covariates. For outcomes with significant main effects, we further employed interaction models to formally test whether these associations were moderated by group (22q11.2DS vs. siblings). All statistical tests were subjected to Benjamini-Hochberg FDR correction separately within each vigilance stage and outcome domain (psychiatric vs cognitive).

### 2.5 Code availability

The complete code base can be found here: https://github.com/liobaberndt/CPNS/tree/paper/2026-tp-22qds#.

## 3 Results

### 3.1 Combined AMPA, NMDA, GABA-A and GABA-B receptor dynamics optimally modelled EEG during wake and sleep states

To identify the optimal combination of glutamatergic and GABAergic receptor types for modelling wake-sleep EEG dynamics, we performed model comparison across models with different combinations of AMPA, NMDA, GABA-A, and GABA-B receptors (figure 1A). The model incorporating all four receptor types (AMPA, NMDA, GABA-A, and GABA-B) emerged as the best explanation of the observed power spectral density when accounting for model complexity, with the lowest BIC value (BIC=24.3). Models including NMDA (ΔBIC = 655.4) or GABA-B (ΔBIC = 549.5) receptors alone showed substantially worse performance compared to the winning model (figure 1B). Additional model comparison metrics presented in the supplementary materials (section 7.3.2) consistently identified the same winning model, further supporting the robustness of this finding.

The winning model implements a canonical cortical microcircuit coupled with thalamic populations (figure 1A). The cortical component comprises four layers with distinct neural populations: superficial pyramidal cells (SP) and interneurons (SI) in layer 2/3, spiny stellate cells (SS) in layer 4, deep pyramidal cells (DP) and interneurons (DI) in layer 5 and thalamic projections (TP) neurons in layer 6. These cortical populations interact with thalamic relay (RL) and reticular (RT) nuclei, completing a thalamocortical circuit. Each connection in this architecture is mediated by specific receptor types, with AMPA and NMDA receptors primarily facilitating excitatory transmission, and GABA-A and GABA-B receptors mediating inhibitory signalling, each operating at distinct timescales: fast AMPA (2.2ms), intermediate GABA-A (5ms), slow NMDA (100ms), and very slow GABA-B (300ms). The model successfully captured the empirical spectral differences between 22q11.2DS carriers and siblings across all vigilance states (figure 2A,B). Variance explained (*r*^2^) was consistently high across all states (figure 2C), with mean *r*^2^ values exceeding 0.95 for both groups.

**Figure 2.**
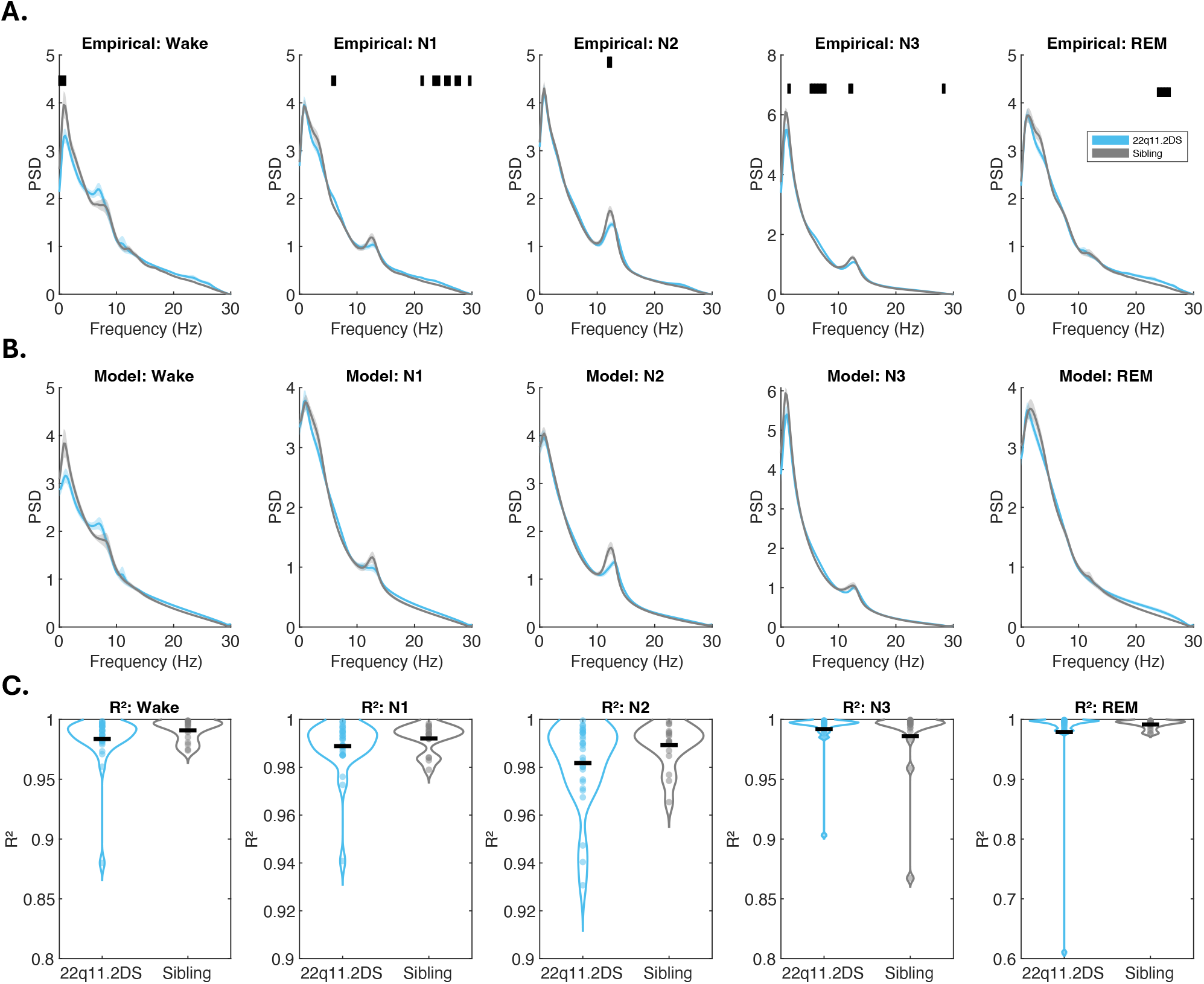
Model Performance Across Sleep Stages. **A.** Actual empirical spectral densities showing group differences (blue = 22q11.2DS; grey = siblings) at the Cz sensor with significant differences (t-test; p < 0.05) marked by black vertical lines. **B**. Model-predicted spectral densities of the winning model. **C**. Variance explained (*r*^2^) by the model for each state, with individual dots representing single participants.

Although the model including all four receptor types provided the best fit to the data, parameter recovery analyses indicated that several parameters were not reliably identifiable. We therefore fixed these non-recoverable parameters to their prior values (denoted as dotted and transparent arrows in figure 1A) and re-estimated the model for all participants. All results reported are based on this reduced, re-estimated model. Full details on parameter recovery and model re-specification are provided in supplementary section 7.3.2.

### 3.2 Systematic Increase in Synaptic Alterations from Wake to REM Sleep in 22q11.2 Deletion Syndrome

We used PEB to assess group-level differences in synaptic parameters across vigilance states. The number of parameters showing significant group differences increased from wakefulness to REM sleep, suggesting that synaptic differences between 22q11.2DS carriers and sibling controls became more widespread in later sleep stages. However, the specific connections and receptor types involved varied across stages and did not form a consistent or easily interpretable pattern. Full stage-specific results and visualisations are provided in supplementary section 7.4.1.

### 3.3 NMDA-R Gain Increase Drives System-Level Alignment between EEG in 22q11.2DS and siblings

We investigated the potential impact of broad receptor-specific pharmacological interventions by systematically decreasing/increasing the gain of each modelled receptor system (AMPA, GABA_A_, GABA_B_, NMDA) across all connections in the model. We evaluated how effectively each manipulation reduced the PSD discrepancy between the 22q11.2DS and sibling control group averages, as measured by the proportional reduction in mean squared error.

Across all vigilance states, increases in NMDA-R gain consistently produced the largest and most reliable improvements in alignment with sibling PSDs. These effects were particularly pronounced during N2 and N3 sleep, yielding effect sizes of 0.32 and 0.45, respectively (figure 3A). Frequency-specific improvements (figure 3B,C) were strongest in the delta and lower theta ranges, with additional gains extending into the spindle frequency range during NREM stages. In lighter sleep stages (N1), REM, and Wake, NMDA-R modulation still yielded significant improvements, albeit with smaller effect sizes (figure 3B,C).

**Figure 3.**
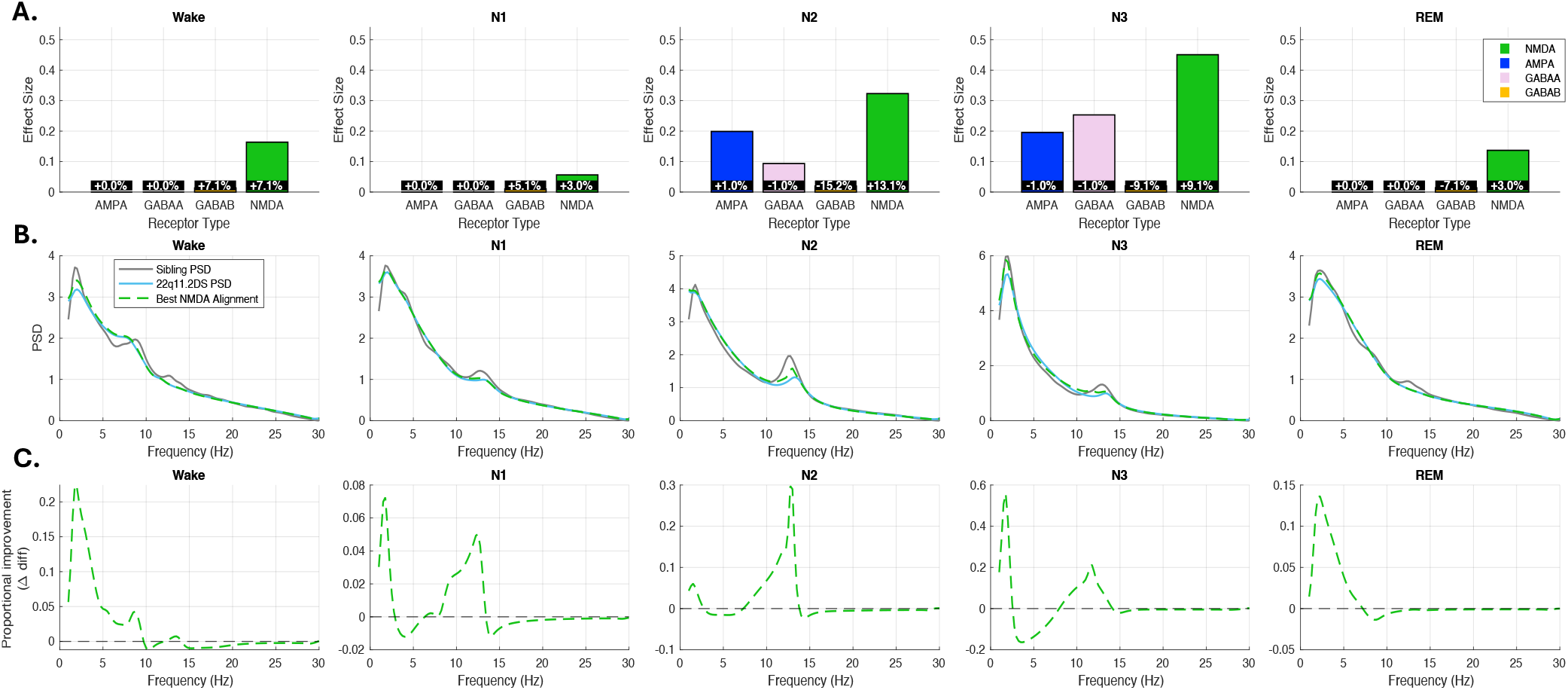
System-wide receptor modulation analysis across vigilance states. **Top row:** Bar plots show the effect sizes for global gain adjustments in each receptor system. Effect size is defined as the proportional reduction in mean squared error between the group-averaged 22q11.2DS PSD and the sibling control PSD following receptor-specific modulation. Each bar reflects the maximum effect size achieved for that receptor system across the tested gain values. The inset box within each bar indicates the specific per gain adjustment that produced the best alignment. Only NMDA-R gain increases produced consistent and substantial improvements across all vigilance states. **Middle row:** Line plots show PSDs for the sibling control group (grey), the 22q11.2DS group (blue), and the best-fitting NMDA gain simulation (green dashed) for each sleep stage. Simulations reflect the NMDA-R gain parameter setting that yielded the lowest MSE relative to the sibling PSD. **Bottom row:** Frequency-specific proportional improvement in alignment (Δ diff), showing the difference between the baseline and NMDA-R-perturbed mismatch with the sibling PSD. Positive values indicate improved alignment. NMDA-R gain increases primarily enhanced alignment in the delta, theta, and alpha frequency ranges during NREM sleep.

To determine which specific pathways contributed to these improvements, we then applied targeted parameter perturbations to isolate the effects of individual NMDA-R-mediated synaptic connections. These analyses revealed that the observed alignment benefits were primarily driven by increased NMDA-R conductance on the self-connections of superficial pyramidal cells, and on the NMDA-R connections from spiny stellate cells to pyramidal cells in the superficial layers (supplementary materials 7.6).

### 3.4 Associations of AMPA-R-Mediated Connectivity and Thalamocortical Delay with Anxiety and Sleep Problems

Regression analysis showed that during wakefulness, stronger AMPA-R-mediated connectivity from superficial pyramidal cells to interneurons was associated with lower anxiety symptoms across groups (main effect: *β* = −0.32, *p*_*FDR*_ = 0.032), with no significant group interaction (figure 4A,B). In N3 (slow-wave) sleep, increased thalamocortical delay was significantly associated with greater sleep problems, and this relationship differed by group (group × parameter interaction: *β* = 0.34, *p*_*FDR*_ = 0.006, figure 4A,B). No other significant associations were observed. All models included age, sex, and group as covariates.

**Figure 4.**
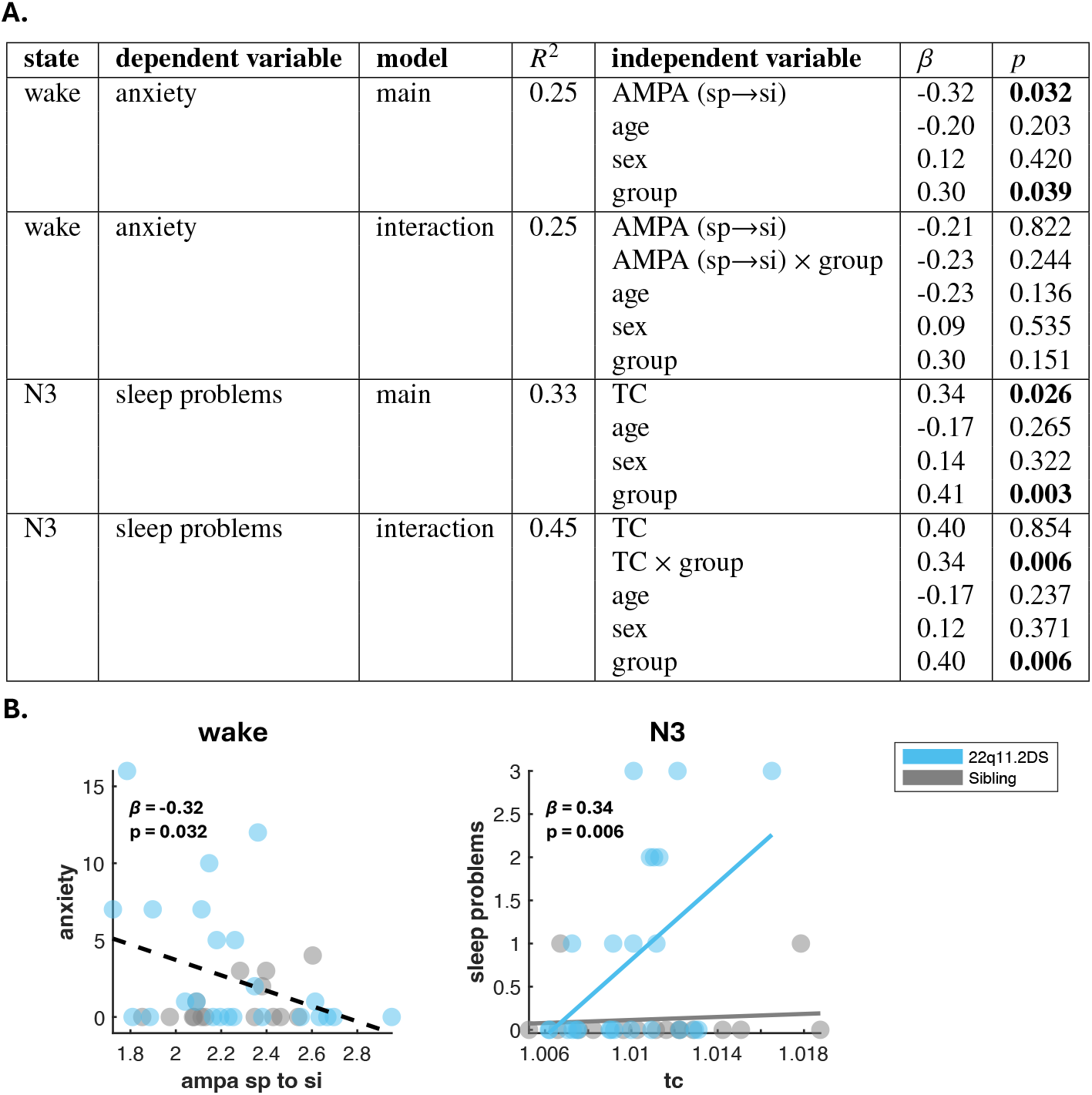
Associations between thalamocortical model parameters and psychiatric symptoms in 22q11.2DS. **A.** Summary table of significant regression results examining the relationships between thalamocortical model parameters and psychiatric or sleep symptoms. For each sleep state and dependent variable, both main effects and interaction (group × parameter) models are shown. The table reports the model *R*^2^, independent variables, standardised beta coefficients (*β*), and *p*-values (*p*). Significant effects (*p*_*FDR*_ < 0.05) are highlighted in bold. **B**. Scatter plots illustrating significant associations. Each point represents an individual, with 22q11.2DS participants shown in blue and sibling controls in grey. When there was a significant group × parameter interaction, separate regression lines were shown for each group (blue for 22q11.2DS, grey for siblings). When there was no significant interaction, a single regression line (black, dashed) was shown for all participants.

## 4 Discussion

In this study, we leveraged computational modelling to infer the synaptic basis of sleep EEG alterations in 22q11.2DS. By systematically perturbing synaptic parameters within a thalamocortical model, we identified alterations that may underlie the observed dynamics. Our central finding was that increasing NMDA receptor gain consistently improved the alignment of 22q11.2DS power spectra with those of neurotypical siblings across all vigilance states, implying NMDA receptor hypofunction as a key driver of altered sleep neurophysiology in 22q11.2DS and highlighting NMDA systems as a potential therapeutic target. In addition to this core result, we found that synaptic alterations became more extensive across sleep stages, and identified associations between specific parameters and psychiatric symptoms, including links between thalamocortical connectivity and sleep problems, and between AMPA-R-related pathways and anxiety.

### 4.1 Sleep As A Potential Amplifier Of Synaptic Dysfunction

Sleep may amplify synaptic dysfunction in 22q11.2DS. We observed that synaptic alterations, which were relatively subtle during wakefulness, became progressively more pronounced across sleep stages. Sleep here could act like a “stress test” for neural circuits in 22q11.2DS. This process appears to unmask underlying vulnerabilities, potentially being linked to the common observation that sleep disturbances are often associated with more severe psychiatric symptoms in 22q11.2DS^7^ and related neurodevelopmental disorders^30,31,32,33,34^. Alternatively, the more regular and persistent nature of network activity during sleep-characterised by stable oscillatory patterns and reduced variability compared to wakefulness-may make underlying synaptic dysfunctions more apparent^35,36,37^.

### 4.2 Therapeutic Potential of Targeting NMDA Receptor Systems

Our receptor-level perturbation analysis identified NMDA-R gain enhancement as the most effective manipulation for shifting 22q11.2DS network dynamics toward control-like patterns, with consistent effects across all vigilance states. These findings support the hypothesis that glutamatergic dysfunction-specifically, NMDA receptor hypofunction-contributes to altered circuit dynamics in 22q11.2DS.

This aligns with evidence for NMDA receptor hypofunction in 22q11.2DS mouse models^38^, a deficit potentially linked to mitochondrial-mediated alterations in intracellular Ca^2+^ handling^39^. While our DCM framework abstracts these microscopic dynamics into a single ‘gain’ parameter, this parameter could potentially capture the functional impact of such Ca^2+^ dysregulation on synaptic signaling. These findings also accord with long-standing models of NMDA dysregulation in schizophrenia and related disorders^40,41,42,43^, and reinforces the potential of NMDA-R-targeting compounds-such as positive allosteric modulators or glycine site agonists-for preclinical investigation^44^. That these effects were most pronounced during NREM sleep suggests a role for NMDA-R-dependent processes in shaping slow-wave activity and sleep-related neural dynamics. Because NMDA-R-gain enhancement improved dynamics in wake, NREM and REM alike, it offers a single, stage-invariant mechanism-far simpler to translate pharmacologically than juggling separate, state-specific interventions.

Finally, our findings demonstrate how in silico pharmacology can extend beyond group-level comparisons to test causal hypotheses about synaptic function. By embedding candidate interventions directly within generative models, this approach offers a mechanistically grounded strategy for identifying targets for future translational and preclinical research.

### 4.3 Altered Thalamocortical Delay and AMPA-Mediated Connectivity are associated with Sleep Disturbances and Anxiety Symptoms

Our findings indicate that sleep problems in 22q11.2DS were most strongly linked to altered thalamocortical delay during N3 (deep non-REM) sleep. Specifically, increased thalamocortical delay was associated with greater sleep problems, and this relationship was significantly moderated by group, suggesting that the impact of thalamocortical dynamics on sleep quality is heightened in 22q11.2DS. This implicates the core thalamocortical system (critical for generating slow waves and maintaining sleep architecture^45,46^) in the subjective experience of poor sleep quality in this population. Coordinated interactions between cortical and thalamic networks underlie non-REM sleep oscillations, with cortical downstates triggering thalamic downstates and subsequent spindle generation^47^. Disruption of this cortico-thalamo-cortical interplay may therefore contribute to the sleep disturbances commonly reported in 22q11.2DS.

In addition, we found that lower anxiety symptoms were associated with stronger AMPA-R-mediated excitation of superficial interneurons during wakefulness, independent of group. This finding suggests that enhanced feedforward inhibition via AMPA receptors may play a protective role in anxiety regulation. However, this interpretation should be considered in light of pharmacological studies reporting anxiolytic effects of AMPA receptor antagonists^48,49^. The mechanisms underlying this discrepancy require further investigation.

## 5 Limitations

First, while our modelling framework aims to incorporate key aspects of biological realism, it remains a simplification and does not include all subcortical structures (i.e., hypothalamic sleep-wake nuclei) or neurotransmitter systems involved in sleep-wake regulation. Within this scope, the model also omits an explicit deep corticoreticular projection, which is potentially relevant for spindle-focused thalamocortical modelling^50^ and should be incorporated in future extensions. More generally, some links remain phenomenological simplifications that do not always map onto anatomical pathways. For example, the spiny-stellate-to-thalamus projection closes the mass-field loop and should not be read as direct corticothalamic wiring from layer 4 and the inhibitory self-connections on excitatory cells also do represent inhibitory autapses onto principal cells. They parameterise net inhibitory conductances on each population’s lumped dynamics and effective recurrent inhibition. In addition, by employing a canonical microcircuit model that includes a granular spiny-stellate layer, we assume a laminar organization that may not precisely match the cytoarchitectural diversity of all cortical regions contributing to the vertex. While this provides a robust effective model of midline cortico-thalamic dynamics, it necessarily abstracts away the fine-scale microanatomical differences between the underlying neural generators. Second, our parameter perturbation approach has inherent constraints. System-wide scaling captures global changes but abstracts away pharmacological specificity, such as receptor subtype selectivity, pharmacokinetics, and off-target effects. Third, the magnitude of simulated changes in PSD resulting from optimal adjustments was consistently small. Although these manipulations reduced the MSE metric, the resulting spectral shifts were subtle, suggesting either limited model sensitivity or that individual parameter changes exert only modest effects. Fourth, our reliance on group-averaged EEG data masks important sources of inter-individual variability within both 22q11.2DS and control groups.

This is particularly significant given the broad age range of our participants (6-20 years), during which the brain undergoes substantial developmental changes. By averaging across this wide age range, we may have obscured important developmental differences in neural dynamics and their modulation, and our findings reflect alignment toward average control patterns rather than personalised trajectories. Fifth, our spectral predictions rely on a frequency-domain linearisation of delays. While this is standard in spectral DCM, Schöbi et al. [51] have shown that approximations of delays in DCM can introduce errors, with accuracy degrading as the product |*ωτ*| grows. With our longest fitted delays (CT: 5-7 ms), this becomes relevant above 23 Hz. Estimates below that threshold are less affected. Our primary finding concerns NMDA receptor gain, where conduction delays (1-7 ms) are small relative to the NMDA time constant (100 ms), placing this inference in the robust regime. Lastly, despite using LASSO regularisation to address the high-dimensional nature of our regression analysis, the relatively small sample size compared to the number of predictors remains a limitation.

## 6 Conclusion

Our findings demonstrate that synaptic circuit alterations in 22q11.2DS are strongly dependent on brain state, with the most pronounced differences emerging during sleep. This state-dependence suggests that static models may miss important aspects of circuit dysfunction and highlights the value of probing neural dynamics across different vigilance states. Motivated by these results, we are now collecting longitudinal EEG and behavioural data to track how synaptic and network changes relate to clinical symptoms over development. In parallel, mouse models of 22q11.2 deletion will allow tests of the causal impact of specific synaptic perturbations on sleep and network dynamics. By pharmacologically manipulating mouse NMDA receptor systems exactly as predicted by our model, we can directly evaluate which interventions restore network activity toward the control regime before advancing to human trials. If the state-dependent circuit alterations observed in humans are recapitulated in mice, this will provide a platform for targeted pharmacological interventions such as NMDA receptor modulation in preclinical models. Ultimately, integrating longitudinal human studies with mechanistic animal work will be essential for translating computational and circuit-level insights into effective, developmentally informed treatments for 22q11.2DS and related neurodevelopmental disorders.

## Data Availability

All data produced are available online at: Nicholas A Donnelly Ullrich Bartsch Hayley A Moulding Christopher Eaton Hugh Marston Jessica H Hall Jeremy Hall Michael J Owen Marianne BM van den Bree Matt W Jones (2022) Sleep EEG in young people with 22q11.2 deletion syndrome: A cross-sectional study of slow-waves, spindles and correlations with memory and neurodevelopmental symptoms eLife 11:e75482.
https://doi.org/10.7554/eLife.75482

## Data availability statement

This study did not generate any new data. All analyses were conducted on previously published data from the IMAGINE-ID/ECHO study, available as part of the original publication by Donnelly et al. (2022)^8^. Source Data files are included with that publication.

## Acknowledgments

We gratefully acknowledge Ullrich Bartsch, Hayley A. Moulding, Christopher Eaton, Hugh Marston, Jessica H. Hall, and Michael J. Owen, whose previous work and data collection made this study possible. We thank the Sleep Detectives team for their valuable input during data analysis, modelling, and manuscript preparation. We are especially grateful to the Lived Experience Advisory Group consisting of families with lived experience of 22q11.2DS and their children, plus support charities (Max Appeal and Unique), for their regular input and feedback throughout the project. Finally, we thank all participants and their families for their invaluable contributions to this research.

## Funding

This work was supported by a Wellcome grant (226709/Z/22/Z).

## Competing interest

The authors declare no competing interests.

## 7 Supplementary Materials

### 7.1 Original study design

This study was originally reported by Donnelly et al. (2022)^8^ and is reproduced here for contextualisation of our modelling approach.

#### 7.1.1 Participants and Recruitment

Participants were recruited through the Experiences of Children with copy number variants (ECHO) study^24^. The study included 28 individuals diagnosed with 22q11.2 Deletion Syndrome and 17 unaffected siblings serving as controls, selected based on proximity in age to the affected participant. Given the exploratory nature of this cross-sectional study involving a rare neurodevelopmental condition, the sample size represented the maximum number of participants who consented to undergo EEG recording during the study period. Confirmation of the 22q11.2 deletion status (presence or absence) was obtained from Medical Genetics laboratory reports or through microarray analysis conducted at the MRC Centre for Neuropsychiatric Genetics and Genomics, Cardiff University. Ethical oversight was provided by the NHS Southeast Wales Research Ethics Committee, which approved all study protocols. Informed consent was obtained from primary carers for all participants. Additionally, participants aged 16 years or older who possessed the capacity to consent provided their own assent. Demographic characteristics can be found in supplementary table 1. Within the 22q11.2DS group, prescribed medications included melatonin (n=4), methylphenidate (n=1), and sertraline (n=1). No control participants were taking psychiatric medications. A history of epilepsy or seizure disorder was an exclusion criterion, and no participants reported such conditions. All data collection, including PSG and cognitive tasks, occurred during single visits to the participants’ homes. These visits were scheduled for participant convenience, typically on weekends or during school holidays. For sibling pairs, data acquisition was completed within the same visit.

**Supplementary table 1.** Demographic and Psychiatric Characteristics. Table shows demographic information, cognitive abilities (FSIQ), psychiatric symptoms, and sleep architecture metrics, with statistical comparisons between 22q11.2DS (n=28) and sibling control (n=17) groups. Adapted from8.

| Variable | Group |  | Type | Statistic<br>(95% CI) | p-value |
| --- | --- | --- | --- | --- | --- |
|  | 22q11.2DS<br>n=28<sup>a</sup> | Sibling<br>n=17<sup>a</sup> |  |  |  |
| Age @ EEG | 14.6 (3.4) | 13.7 (3.4) | Diff<sup>b</sup> | 0.90 [-1.22, 3.01] | 0.397 |
| Sex | | | $\chi^2$<sup>c</sup> | 0 | 1 |
| Female | 14 (50%) | 9 (53%) |  |  |  |
| Male | 14 (50%) | 8 (47%) |  |  |  |
| Sleep Problem | 1.32 (1.70) | 0.24 (0.56) | OR<sup>d</sup> | 6.27 [2.12, 18.56] | 0.001 |
| FSIQ | 76 (13) | 105 (27) | Diff<sup>e</sup> | -28.70 [-40.48, -16.92] | <0.001 |
| missing | 0 | 1 |  |  |  |
| Anxiety Sx | 5.0 (7.8) | 1.4 (2.8) | OR<sup>d</sup> | 3.10 [1.93, 4.99] | <0.001 |
| ADHD Sx | 6.0 (6.0) | 0.7 (2.1) | OR<sup>d</sup> | 9.46 [5.12, 17.48] | <0.001 |
| ASD Sx | 11 (6) | 1 (2) | OR<sup>d</sup> | 7.46 [4.76, 11.70] | <0.001 |
| missing | 1 | 1 |  |  |  |
| Psychotic Exp |  |  | OR<sup>f</sup> | 4.05 [0.70, 43.67] | 0.096 |
| No PE | 18 (64%) | 15 (88%) |  |  |  |
| PE | 10 (36%) | 2 (12%) |  |  |  |
| N1 (%) | 10.4 (4.7) | 13.6 (4.3) | Diff<sup>e</sup> | -2.71 [-5.05, -0.36] | 0.044 |
| N2 (%) | 26.2 (8.2) | 27.1 (5.9) | Diff<sup>e</sup> | -1.09 [-5.15, 2.97] | 0.620 |
| N3 (%) | 30 (7) | 25 (6) | Diff<sup>e</sup> | 5.47 [1.98, 8.96] | 0.009 |
| REM (%) | 14.4 (4.6) | 18.2 (5.6) | Diff<sup>e</sup> | -4.20 [-7.10, -1.30] | 0.012 |
| N1 Lat (min) | 23 (18) | 21 (9) | Diff<sup>e</sup> | 3.49 [-5.54, 12.51] | 0.470 |
| REM Lat (min) | 143 (69) | 140 (49) | Diff<sup>e</sup> | 9.37 [-19.31, 38.05] | 0.549 |
| Sleep Eff (%) | 88 (8) | 89 (9) | Diff<sup>e</sup> | -1.85 [-5.83, 2.14] | 0.398 |
| TST (min) | 456 (122) | 485 (79) | Diff<sup>e</sup> | -27.21 [-88.49, 34.08] | 0.413 |
| Awakenings | 42 (52) | 42 (40) | Diff<sup>e</sup> | 3.10 [-19.73, 25.93] | 0.802 |

<sup>a</sup> Mean (SD); n (%)
<sup>b</sup> Linear Model; <sup>c</sup> $\chi^2$ Test; <sup>d</sup> GLMM; <sup>e</sup> LMM; <sup>f</sup> Fisher's Test
Sx = Symptoms; Exp = Experiences; Lat = Latency; Eff = Efficiency; TST = Total Sleep Time

#### 7.1.2 Psychiatric and Cognitive Assessment

Psychopathology symptoms and subjective sleep quality were evaluated using the Child and Adolescent Psychiatric Assessment (CAPA) interview^52^, administered to either the participant or their primary carer during the home visit. Symptoms related to Autism Spectrum Disorder were screened via the Social Communication Questionnaire (SCQ), completed by the primary carer. Cognitive ability, specifically Full-Scale IQ (FSIQ), was assessed using the Wechsler Abbreviated Scale of Intelligence (WASI)^53^.

#### 7.1.3 CANTAB Cognitive Assessment

Cognitive function was assessed using the Cambridge Neuropsychological Test Automated Battery (CANTAB)^28^, a computerised set of neuropsychological tests administered on a touchscreen device. The following tests were administered:

##### Working Memory and Processing Speed

Working memory capacity was assessed using the Spatial Working Memory (SWM) task^54^, which requires participants to search for tokens hidden in boxes while remembering and avoiding previously searched locations. Processing speed was measured through reaction times across various CANTAB tasks.

##### Sustained Attention

The Rapid Visual Processing (RVP)^55^ task was used to assess sustained attention. Participants were required to detect target sequences of digits in a continuous stream of numbers. Performance was quantified using the RVP A’ score, a signal detection measure that accounts for both hits and false alarms, providing a measure of sensitivity to the target sequence independent of response tendency.

##### Planning Ability

Planning ability was assessed using the Stockings of Cambridge (SOC) task^54^, a computerised version of the Tower of London test. Participants were required to move colored balls in a specified minimum number of moves to match a target arrangement. Three measures were derived:

- SOC Initial Time: Planning time before the first move
- SOC Subsequent Time: Average time for moves after the first move
- SOC Problems Solved: Total number of problems solved in the minimum number of moves

All tests were administered according to standardised CANTAB protocols, with task difficulty increasing progressively based on participant performance. Raw scores were converted to standardised scores based on age-appropriate normative data provided by CANTAB.

#### 7.1.4 Wisconsin Card Sorting Test

The Wisconsin Card Sorting Test (WCST)^56^ was administered as an additional measure of executive function, specifically cognitive flexibility and set-shifting abilities. In this computerised version of the test, participants were required to sort cards according to different rules (colour, shape, or number) that changed without warning. The task required participants to identify the current sorting rule through trial-and-error learning and adapt their responses when the rule changed. Performance was measured through several key indices:

- Perseverative Errors: Number of times participants persisted with an incorrect sorting rule despite negative feedback
- Non-perseverative Errors: Number of other types of incorrect responses
- Categories Completed: Number of times participants successfully identified and maintained the correct sorting rule

These measures provided a comprehensive assessment of cognitive flexibility, rule-learning abilities, and the capacity to adapt to changing task demands.

#### 7.1.5 Polysomnography (PSG) Data Acquisition

Overnight PSG recordings were conducted in participants’ homes using ambulatory systems to minimise disruption to normal sleep routines. High-density EEG was acquired using a 60-channel Geodesic Net (EGI), connected to a BE Plus LTM amplifier running Galileo software (EBNeuro). Standard PSG signals, including EOG, EMG, ECG, respiratory effort (inductance plethysmography), oxygen saturation (pulse oximetry), and nasal airflow, were recorded using an Embla Titanium amplifier. All signals were digitised at a sampling rate of 512 Hz with an online high-pass filter setting of 0.1 Hz. The EEG was referenced online to the Cz electrode. Participants slept in their own beds and could move freely during the night.

#### 7.1.6 Sleep Scoring

Sleep stages were visually scored in 30-second epochs by an experienced technician based on standard AASM criteria, using a montage comprising 6 EEG channels and all auxiliary PSG channels. Epochs containing significant artefact were excluded from subsequent EEG analyses. Standard sleep architecture variables were derived, including total sleep time, sleep efficiency, latencies to N1 and REM sleep, and the percentage of total sleep time spent in N1, N2, N3, and REM sleep stages.

#### 7.1.7 EEG Data Processing

EEG data processing relied on established methodologies implemented in MATLAB (Mathworks, Natick, MA, USA), utilising custom scripts and functions from the EEGLAB toolbox^57^ where appropriate. Raw EEG data were imported into EEGLAB. The pre-processing pipeline included downsampling to 128 Hz, high-pass filtering at 0.25 Hz, removal of 50 Hz line noise using the PREP plugin^58^, suppression of artefacts using Artifact Subspace Reconstruction^59^ via the clean_rawdata plugin, and re-referencing to the common average. Independent Component Analysis (ICA) was performed using the AMICA plugin^60^, and components identified as non-brain activity (e.g., ECG, EMG, EOG) by the ICLabel algorithm were removed^61^. An additional automated epoch-level artefact rejection procedure was applied to N2 and N3 sleep epochs combined, using iterative thresholding based on power in specific frequency bands (beta: 16–25 Hz; delta: 1–4 Hz) relative to surrounding epochs, detection of clipped signals, and outlier detection based on signal Root Mean Square (RMS) and Hjorth parameters^62,63^. This channel-wise procedure did not result in significant group differences in the proportion of N2 or N3 data removed. Epochs identified as artefact during the manual sleep scoring process were also marked for exclusion.

Spectrograms representing time-varying power across the night were calculated using the multitaper method (30s window, 10s step, 1 Hz bandwidth)^64^. Power spectral density (PSD) for frequencies between 0.25 and 30 Hz was computed for each epoch using Welch’s periodogram method (4s Hanning window, 2s step) and expressed in decibels (dB). This calculation was performed on both the original and z-scored EEG signals. To separate oscillatory and aperiodic signal components, the Irregular Resampling Auto-Spectral Analysis (IRASA) method was applied^65^. Group differences in PSD at each frequency were statistically assessed using linear mixed models, with cluster-based permutation tests applied to correct for multiple comparisons across frequencies^66^.

### 7.2 EEG spectral power at Cz Electrode across sleep stages for 22q11.DS and their siblings

Donnelly et al. (2022)^8^ reported the most pronounced differences in spectral power between 22q11.2DS individuals and their siblings at the Cz electrode. These differences are illustrated in supplementary figure 1 and lay the foundation for our subsequent modelling.

**Fig S1.**
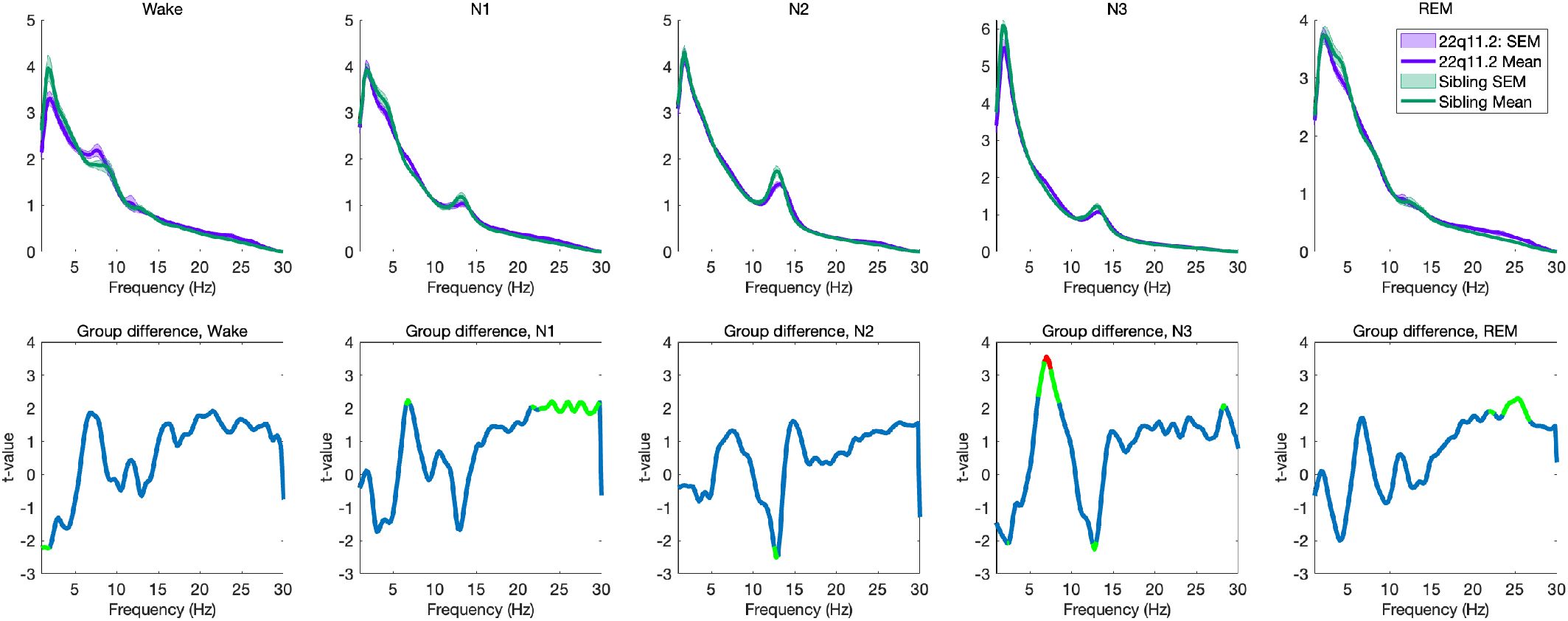
EEG Spectral Power at Cz Electrode Across Sleep Stages. The Cz electrode showed the most pronounced group differences in power spectral density (PSD), as previously reported by^8^. **Top row:** Mean spectral power (±SEM) at the Cz electrode for individuals with 22q11.2 deletion syndrome (purple) and their sibling controls (green) across Wake, N1, N2, N3, and REM sleep stages. The x-axis shows frequency (Hz), and the y-axis shows power (*μV*^2^ / Hz, log scale). **Bottom row:** T-values indicating group differences in spectral power across frequencies. Positive t-values (above zero) reflect greater power in the 22q11.2 group, while negative t-values (below zero) indicate reduced power relative to sibling controls.

### 7.3 Thalamo-Cortical-Modelling

We employed Dynamic Causal Modelling^16^ to analyse spectral densities in EEG signals. DCM utilises parameterised dynamical systems models, specifically state-space formulations of differential equations—so-called neural mass models—which describe how variables of interest (e.g., membrane potentials) evolve as a function of connectivity parameters^16^. We utilised the thalamo-cortical DCM variant developed by Shaw et al. (2020)^19^, which explicitly models interactions between cortical layers and thalamic nuclei.

#### 7.3.1 Modelling procedure

##### Model Architecture

The thalamo-cortical model incorporates interacting, layer-resolved cortical and thalamic populations, following architectures described by Dougles and Martin (2004)^67^, and Gilbert and Wiesel (1983)^68^. The model is based on conductance-based equations inspired by the Hodgkin–Huxley and Morris–Lecar formalisms^69^. It includes pyramidal and interneuron populations in cortical layers 2/3 and 5, stellate cells in layer 4, thalamic-projection pyramidal cells in layer 6, and thalamic relay and reticular populations^70^.

Population dynamics are governed by the following set of coupled differential equations:

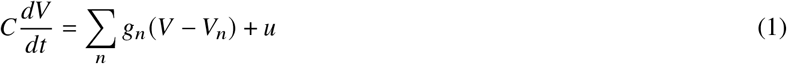

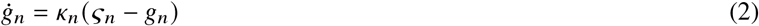

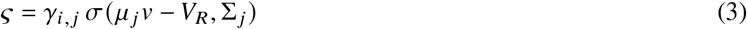

Here, *V* represents the membrane potential, *g*_*n*_ denotes conductance due to receptor *n*, and *V*_*n*_ is the corresponding reversal potential. *C* is the membrane capacitance, and *u* represents external or internal input currents. *k*_*n*_ is the decay rate of channel *n, γ*_*i, j*_ the coupling parameter between populations *i* and *j*, and *σ* is a sigmoid function converting membrane potential into population firing rates.

The model includes AMPA, NMDA, GABA-A, GABA-B, M-type, and H-type conductance channels. M and H channels are specific to layer 6 thalamic-projecting pyramidal cells and thalamic relay populations, facilitating their characteristic bursting behaviour. Building on Moran et al.s NMDA model^71^, we incorporate a voltage-dependent magnesium block for NMDA receptors:

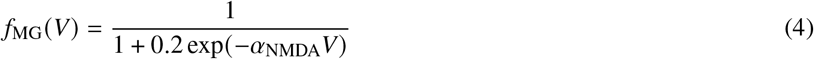

Reversal potentials and decay rates were derived from established literature to ensure physiological accuracy^72,73,74^. M-channels influence intrinsic membrane dynamics and the excitation–inhibition balance, while H-channels affect resting membrane potential and network oscillations^75,76,77,78^. GABA-B receptors contribute to macroscale oscillations, particularly in the gamma range^79,80^.

##### Generative Model Specification

Following standard spectral DCM approaches, we computed the power spectrum using the Laplace transform. This frequency-domain linearisation differs from the approach of^19^, who performed time-domain numerical integration (Euler method with delays) followed by FFT. After performing a local linearisation by numerically computing the system’s Jacobian about a fixed point, we derived the transfer function from this linearised approximation. The Laplace transform at frequency i, in hertz, is given by:

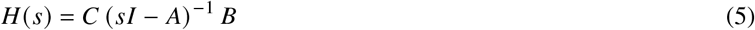

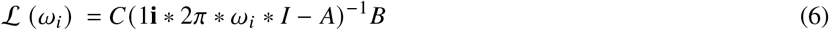

Here, equation (5) represents the “canonical form” Laplace transform, where *H(s)* is the transfer function and *s* is the complex Laplace variable. Equation (6) shows the Laplace transform evaluated at specific frequency *w(i)* in hertz. *A* represents the system Jacobian matrix (*df/dx*), *B* the input Jacobian (*df/du*), and *C* a vector of weights determining each state element’s contribution to the output potential at Cz. Accordingly, the signal at Cz is modelled as a weighted sum of population membrane potentials (after linearisation), with these weights estimated during model inversion. This transfer function approach offers computational efficiency compared to numerical integration of model equations. Unlike conventional spectral DCM, we did not explicitly include spectral noise components such as 1/f “aperiodic” components or a discrete cosine basis set for frequency-domain neuronal fluctuations. Instead, the model output is given by the Laplace transform of the linearised system, evaluated in the frequency domain without numerical integration of stochastic processes. Exogenous input is applied only to the thalamic relay and reticular populations, and is characterised by a flat (white) power spectrum.

##### Model Optimisation and Free Energy

Model parameters were optimised using variational Bayes to maximise the free energy^81^, which provides a lower bound on the log model evidence. The free energy *F* is defined as:

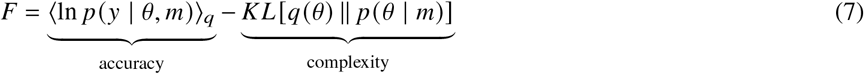

Here, *p* (*y* | *θ, m*) denotes the likelihood of the observed data *y* given parameters *θ* under model *m*, and *q* (*θ*) is the approximate posterior distribution over parameters. The term *KL*[*q*(*θ*) *p* (*θ* | *m*)] is the Kullback–Leibler divergence^82^ between the approximate posterior and the prior distribution *p* (*θ* | *m*).

This framework offers a principled trade-off between model accuracy and complexity^16^, mitigating overfitting while ensuring that the model captures essential features of the observed spectra.

Optimisation was performed using an Expectation-Maximisation scheme for each individual, which iteratively updates the posterior distributions of parameters until convergence. This yields both optimal parameter estimates and their associated uncertainties, enabling robust statistical inference about group differences between individuals with 22q11.2DS and their siblings.

##### Model priors

#### 7.3.2. Model Comparison

##### Model development

The thalamo-cortical model, first introduced by Shaw et al. (2020)^19^ and implemented in this study, represents the culmination of decades of neural–mass model work that has progressively incorporated greater anatomical and physiological detail in order to capture the full neural spectrum. In order to provide context and explain why such model complexity is necessary for investigating sleep neurophysiology, we outline the historical development of neural mass models.

The seminal equations of Wilson and Cowan (1972)^83^ described interacting excitatory (E) and inhibitory (I) populations and established the mathematical foundations of neural-mass modelling. Although highly abstract, these E–I models demonstrated how macroscopic oscillations can emerge from coupled cortical populations. Jansen and Rit (1995)^84^ extended the E–I framework to a cortical column comprising pyramidal cells, excitatory interneurones and inhibitory interneurones. With distinct synaptic time-constants this three-population model reproduced the *α*-band oscillations that dominate wakeful EEG. Dynamic Causal Modelling embedded these neural equations in a Bayesian inversion framework^16^, allowing synaptic parameters to be estimated directly from empirical data. To account for laminar organisation and pharmacological manipulations, the canonical microcircuit (CMC) introduced superficial and deep pyramidal layers together with receptor-specific AMPA, NMDA and GABA_A_ conductances^85,86^. This architecture successfully captured faster *β*/*γ* activity and permitted quantitative assays of drug actions on glutamatergic and GABAergic signalling^87^.

Purely cortical models, however, cannot reproduce oscillations that depend on thalamo-cortical loops and slow inhibitory kinetics. To overcome these limitations, Shaw et al. (2020)^19^ augmented the CMC with thalamic relay (RL) and reticular (RT) nuclei plus corticothalamic projection neurones, yielding an eight-population circuit (five cortical, three thalamic). Critically, the TCM incorporates slow GABA_B_ inhibition as well as intrinsic *M*- and *H*-type currents, features that are essential for reproducing sleep-stage–specific EEG, including spindles and slow-wave activity. Because our research question concerns synaptic mechanisms of sleep in 22q11.2DS—a condition where both cortical and thalamic circuitry are implicated—the TCM provides the minimum biological detail required. Accordingly, the Bayesian model comparison and parameter-recovery analyses reported below are conducted within this thalamo-cortical model family.

##### Nested Model Construction

To systematically evaluate the contribution of different synaptic receptor systems in this thalamo-cortical network, we constructed 15 nested models with varying synaptic connectivity patterns (supplementary figure 2). Each model represents a hypothesis about the essential receptor-mediated connections required to generate sleep-stage specific EEG patterns. The models were constructed by systematically including or excluding connections mediated by four key synaptic receptor types (AMPA, NMDA, GABA-A, GABA-B) between eight key regions: Superficial Pyramidal cells (SP), Superficial Interneurons (SI), Spiny Stellate cells (SS), Deep Pyramidal cells (DP), Deep Interneurons (DI), Thalamic Projection neurons (TP), Thalamic Relay (RL), and Reticular nucleus (RT).

**Fig S2.**
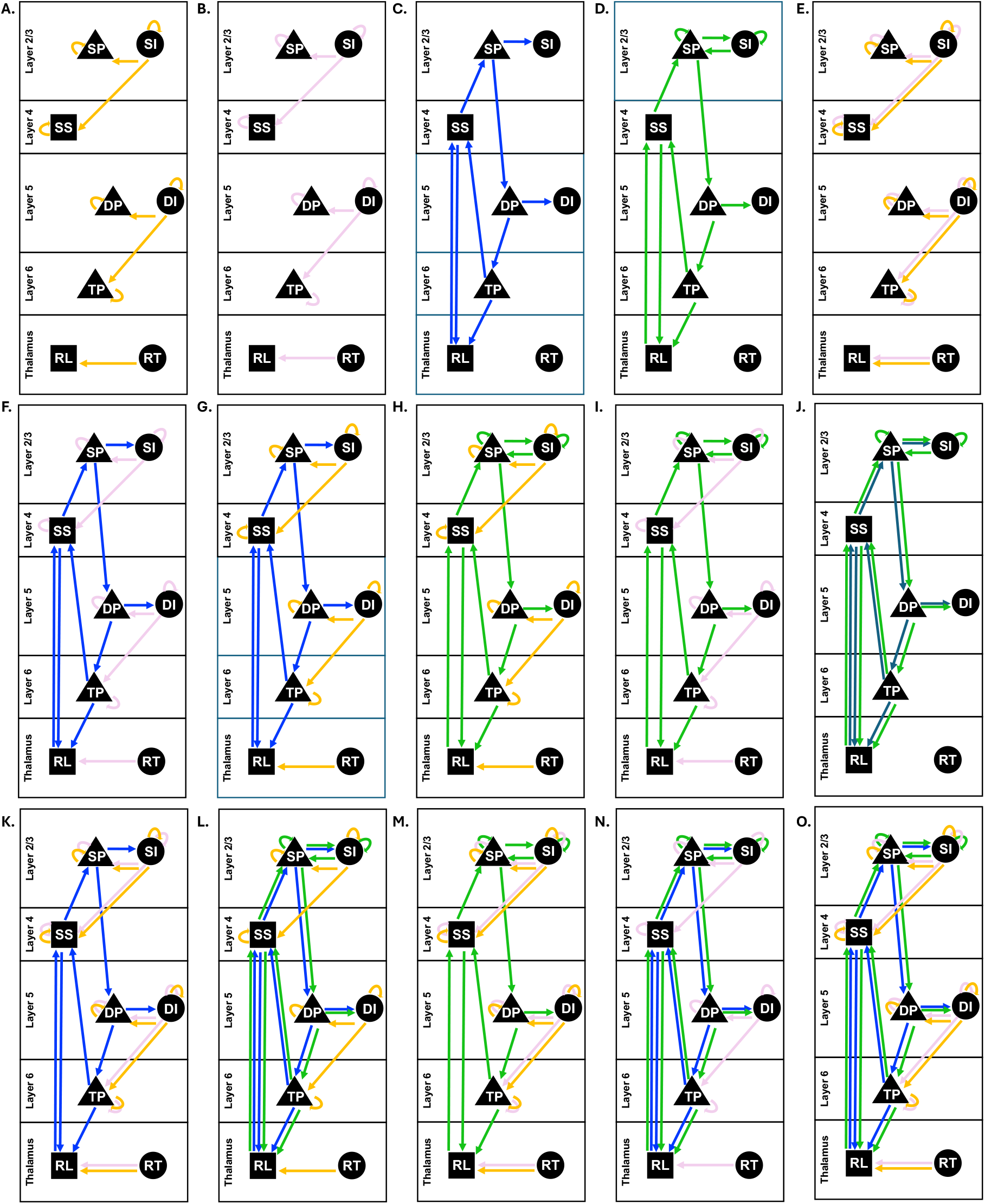
Nested Models of Receptor-Specific Connectivity. Schematic representation of 15 nested dynamic causal models (A-O) showing different receptor-mediated connectivity patterns in the thalamo-cortical network. Each panel represents a distinct model with specific synaptic connections. Regions are organised in anatomical layers: Layer 2/3, Layer 4, Layer 5, Layer 6, and Thalamus with different neural populations: SP (Superficial Pyramidal), SI (Superficial Interneurons), SS (Spiny Stellate), DP (Deep Pyramidal), DI (Deep Interneurons), TP (Thalamic Projection), RL (Relay), and RT (Reticular nucleus). Arrows indicate directed connections, with colours representing different receptor types: blue: AMPA receptors (fast ionotropic glutamate receptors); pink: GABA-A receptors (fast ionotropic GABA receptors); yellow: GABA-B receptors (slow metabotropic GABA receptors); green: NMDA receptors (slow ionotropic glutamate receptors).

##### Bayesian Model Comparison of Receptor-Mediated Connections

Having established our 15 nested models of increasing complexity, we next performed a systematic Bayesian model comparison to determine which combination of receptor-mediated connections best explains the observed sleep EEG data.

To quantitatively evaluate these models, we employed four complementary metrics:

1. **Variational Free Energy (F)**: A lower bound on the log model evidence that balances model accuracy with complexity:

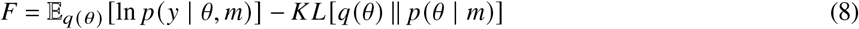

where *q* (*θ*) is the approximate posterior, *p* (*θ* | *m*) is the prior, and *p* (*y* | *θ, m*) is the likelihood. Higher (i.e., less negative) free energy values indicate better model evidence by favouring accurate yet parsimonious models.
2. **Bayesian Information Criterion (iBIC)**: A complexity-penalised criterion for model comparison, computed as:

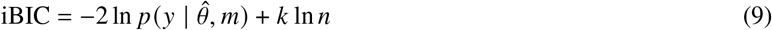

where 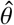 denotes the maximum a posteriori estimate of the parameters, *k* is the number of free parameters, and *n* is the number of observations. Lower iBIC values indicate better model fits. **Posterior Model Probability**: The probability of each model *m* given the data *y*, calculated using softmax over the free energies of all candidate models:

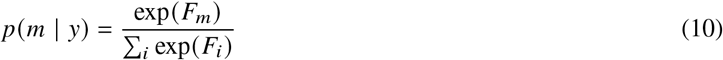

where *F*_*m*_ is the free energy of model *m*. This provides a probabilistic ranking of models.
3. **Log Bayes Factors (lnBF)**: The logarithmic difference in model evidence between each model and the best-performing (winning) model:

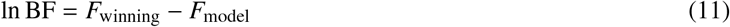

Our analysis revealed a clear hierarchy in model evidence (supplementary figure 3). The model incorporating all four receptor types (AMPA, GABA-A, GABA-B, NMDA) showed the highest posterior probability (0.758) and highest free energy (-12.2). This model also achieved the lowest iBIC (24.3), indicating it provides the optimal balance between model fit and complexity. Notably, the difference in model evidence between the full model and the model without NMDA receptors was small (log Bayes factor < 3), indicating that this does not constitute strong evidence in favour of the more complex model, and that both models provide a comparably good account of the data.

**Fig S3.**
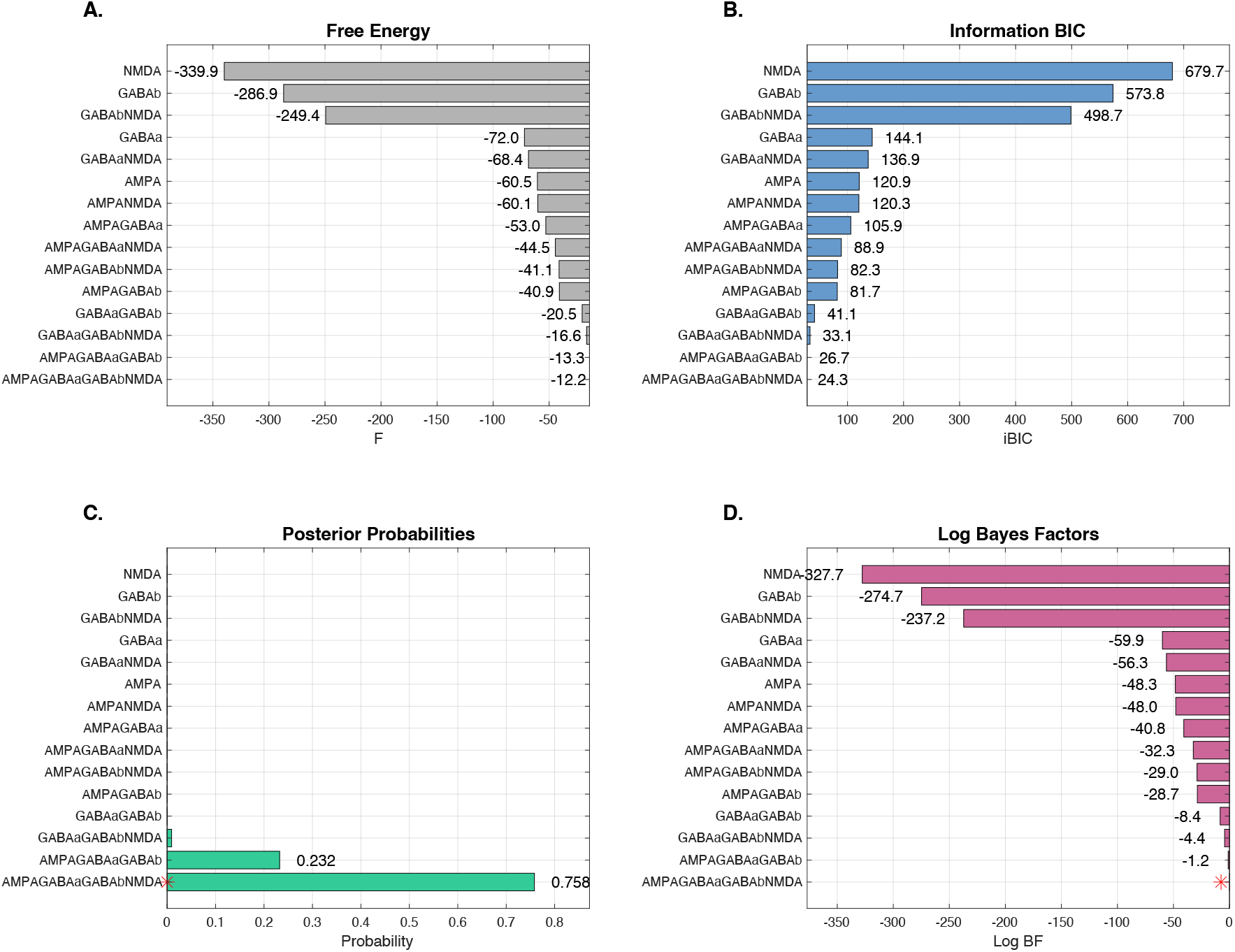
Bayesian Model Comparison Metrics. Comparison of 15 nested models with different combinations of receptor-mediated connections using four Bayesian model comparison metrics. **(A)** Free energy (F) values for each model, where higher (less negative) values indicate better model evidence. **(B)** Information Bayesian Information Criterion (iBIC) scores, with lower values indicating better model parsimony. The full model achieves the lowest iBIC (24.3), suggesting optimal balance between accuracy and complexity. **(C)** Posterior probabilities showing the relative likelihood of each model given the data, with the full model achieving the highest probability of 0.758. **(D)** Log Bayes Factors comparing each model against the winning model, where more negative values indicate stronger evidence against that model. Model names indicate included receptor types. The progressive improvement in metrics with additional receptor types suggests that each receptor system contributes meaningfully to generating sleep-stage specific EEG patterns.

Notably, models with single receptor types showed poor performance, with NMDA-only and GABA-B-only models showing particularly low free energy values (-339.9 and -286.9 respectively) and high iBIC scores (679.7 and 573.8). This indicates that no single receptor type can adequately explain the complex dynamics of sleep-stage transitions. The progressive improvement in model evidence metrics as receptor types are combined suggests that each receptor system plays a distinct and necessary role in generating sleep-stage specific EEG patterns.

#### 7.3.3 Parameter Recovery

To ensure reliable parameter estimation and validate our model’s ability to recover true parameter values, we performed parameter recovery analyses. This involved generating synthetic data from known parameter values and then fitting our thalamo-cortical model to these data to assess recovery accuracy.

We simulated EEG spectra using parameter values drawn from physiologically plausible ranges, then fitted the model to these synthetic data using the same optimization procedures as applied to empirical data. Parameter recovery was quantified using Intraclass Correlation Coefficients (ICC) between true and recovered parameter values, providing a measure of estimation reliability across the parameter space.

To maximize the number of interpretable parameters while ensuring estimation reliability, we employed an iterative parameter fixing strategy. In the first iteration, we identified parameters that remained at their prior values across participants, indicating they were not contributing to model fit, and fixed these at their prior values. We then re-ran parameter recovery to assess how ICC values changed. In the second iteration, we fixed only parameters with ICC values below 0.2, representing the most unreliable estimates, and again re-ran parameter recovery. Finally, in the third iteration, we assessed whether any remaining parameters had ICC values below 0.4^88^, fixing these. This iterative approach prioritizes retaining as many parameters as possible for biological interpretation while ensuring that only well-constrained parameters contribute to our group-level inferences, reducing the risk of spurious findings while maintaining the model’s ability to capture meaningful neurophysiological differences. This procedure resulted in ICC values > 0.4 for all retained parameters (supplementary table 3), with many of these parameters reaching ICC values > 0.70, placing them in the good-excellent range (as suggested by Fleiss (2011)^88^). Parameters were fixed at their prior value (see supplementary table 2) and were taken from Shaw et al. [19]. Model comparison was performed on all models before fixing non-recoverable parameters.

**Supplementary table 2.** Prior means and variances for thalamo-cortical and cortical-cortical synaptic parameters across different receptor types (AMPA, GABA-A, NMDA, and GABA-B) used during model fitting. Additional estimated parameters included J(1:8) (weights mixing the eight population membrane potentials into the channel prediction), and d(1) (spectral smoothing of that prediction), all other parameters were fixed.

| Name | Prior mean | Prior variance |
| --- | --- | --- |
| AMPA_ss_to_sp | 2 | 1/8 |
| AMPA_ss_to_rl | 2 | 1/8 |
| AMPA_sp_to_si | 2 | 1/8 |
| AMPA_sp_to_dp | 2 | 1/8 |
| AMPA_dp_to_di | 2 | 1/8 |
| AMPA_dp_to_tp | 2 | 1/8 |
| AMPA_tp_to_ss | 2 | 1/8 |
| AMPA_tp_to_rl | 2 | 1/8 |
| AMPA_rl_to_ss | 2 | 1/8 |
| GABAA_ss_to_ss | 8 | 1/8 |
| GABAA_sp_to_sp | 18 | 1/8 |
| GABAA_si_to_ss | 10 | 1/8 |
| GABAA_si_to_sp | 10 | 1/8 |
| GABAA_si_to_si | 10 | 1/8 |
| GABAA_dp_to_dp | 8 | 1/8 |
| GABAA_di_to_dp | 6 | 1/8 |
| GABAA_di_to_di | 14 | 1/8 |
| GABAA_di_to_tp | 6 | 1/8 |
| GABAA_tp_to_tp | 8 | 1/8 |
| GABAA_rt_to_rl | 8 | 1/8 |
| NMDA_ss_to_sp | 2 | 1/8 |
| NMDA_ss_to_rl | 2 | 1/8 |
| NMDA_sp_to_sp | 2 | 1/8 |
| NMDA_sp_to_si | 2 | 1/8 |
| NMDA_sp_to_dp | 2 | 1/8 |
| NMDA_si_to_sp | 2 | 1/8 |
| NMDA_si_to_si | 2 | 1/8 |
| NMDA_dp_to_di | 2 | 1/8 |
| NMDA_dp_to_tp | 2 | 1/8 |
| NMDA_tp_to_ss | 2 | 1/8 |
| NMDA_tp_to_rl | 2 | 1/8 |
| NMDA_rl_to_ss | 2 | 1/8 |
| GABAB_ss_to_ss | 8 | 1/8 |
| GABAB_sp_to_sp | 18 | 1/8 |
| GABAB_si_to_ss | 10 | 1/8 |
| GABAB_si_to_sp | 10 | 1/8 |
| GABAB_si_to_si | 10 | 1/8 |
| GABAB_dp_to_dp | 8 | 1/8 |
| GABAB_di_to_dp | 6 | 1/8 |
| GABAB_di_to_di | 14 | 1/8 |
| GABAB_di_to_tp | 6 | 1/8 |
| GABAB_tp_to_tp | 8 | 1/8 |
| GABAB_rt_to_rl | 8 | 1/8 |
| Thalamo-Cortical delay | 3 ms | 1/8 |
| Cortico-Thalamic Delay | 8 ms | 1/8 |
| scale_NMDA | 1 | 1/8 |
| Electrode Gain | 1 | 1/8 |

**Supplementary table 3.** ICC for parameter recovery analysis. Parameters with ICC values < 0.4 were fixed at their prior values. All further analysis were performed on this reduced model.

| Parameter | ICC |
| --- | --- |
| **AMPA** |  |
| AMPA_ss_to_sp | 0.807 |
| AMPA_ss_to_rl | 0.936 |
| AMPA_sp_to_si | 0.800 |
| AMPA_sp_to_dp | 0.528 |
| AMPA_dp_to_di | Fixed |
| AMPA_dp_to_tp | 0.791 |
| AMPA_tp_to_ss | Fixed |
| AMPA_tp_to_rl | Fixed |
| AMPA_rl_to_ss | 0.891 |
| **GABA-A** |  |
| GABA-A_ss_to_ss | 0.779 |
| GABA-A_sp_to_sp | 0.825 |
| GABA-A_si_to_ss | Fixed |
| GABA-A_si_to_sp | 0.467 |
| GABA-A_si_to_si | 0.982 |
| GABA-A_dp_to_dp | 0.650 |
| GABA-A_di_to_dp | 0.706 |
| GABA-A_di_to_di | 0.776 |
| GABA-A_di_to_tp | 0.872 |
| GABA-A_tp_to_tp | 0.836 |
| GABA-A_rt_to_rl | 0.586 |
| **NMDA** |  |
| NMDA_ss_to_sp | 0.731 |
| NMDA_ss_to_rl | Fixed |
| NMDA_sp_to_sp | 0.847 |
| NMDA_sp_to_si | 0.613 |
| NMDA_sp_to_dp | 0.750 |
| NMDA_si_to_sp | 0.636 |
| NMDA_si_to_si | Fixed |
| NMDA_dp_to_di | Fixed |
| NMDA_dp_to_tp | Fixed |
| NMDA_tp_to_ss | Fixed |
| NMDA_tp_to_rl | Fixed |
| NMDA_rl_to_ss | 0.650 |
| **GABA-B** |  |
| GABA-B_ss_to_ss | 0.940 |
| GABA-B_sp_to_sp | 0.903 |
| GABA-B_si_to_ss | Fixed |
| GABA-B_si_to_sp | 0.769 |
| GABA-B_si_to_si | 0.890 |
| GABA-B_dp_to_dp | 0.896 |
| GABA-B_di_to_dp | Fixed |
| GABA-B_di_to_di | Fixed |
| GABA-B_di_to_tp | Fixed |
| GABA-B_tp_to_tp | 0.583 |
| GABA-B_rt_to_rl | 0.903 |
| **Other** |  |
| TC | 0.891 |
| CT | 0.855 |
| scale_NMDA | 0.820 |

### 7.4 Parametric Empirical Bayes Analysis of Group Differences

Following the initial model comparison which established the optimal thalamocortical architecture common to all subjects, we employed Parametric Empirical Bayes (PEB; spm_dcm_peb)^25,26^ to specifically investigate differences in synaptic parameters within this winning model structure between the 22q11.2DS and control groups. PEB provides a hierarchical Bayesian framework, analogous to a General Linear Model, allowing for robust estimation of group effects on the DCM parameters.

A key advantage of PEB over classical statistical approaches, such as applying independent t-tests to each parameter, lies in its principled handling of estimation uncertainty. DCM parameter estimates are inherently associated with a degree of precision (or uncertainty). PEB explicitly incorporates this uncertainty by weighting the contribution of each subject’s parameter estimate to the group-level inference by its posterior precision (inverse variance). Parameters estimated with lower confidence (higher variance) appropriately exert less influence on the group results, whereas classical tests typically ignore this crucial information. This makes PEB particularly robust for analysing complex biophysical models like DCM, where parameters may be estimated with varying degrees of confidence^25^.

To further enhance robustness and identify the most parsimonious explanation for group differences, the PEB analysis pipeline incorporates automatic Bayesian Model Reduction (BMR) and Averaging (BMA; spm_dcm_peb_bmc)^25,89^. BMR systematically compares nested models where specific parameters (or combinations) representing group differences are iteratively ‘switched off’. It evaluates the evidence for each reduced model, effectively pruning away parameters that do not contribute meaningfully to explaining between-subject or between-group variability. Subsequently, Bayesian Model Averaging computes weighted averages of the parameter estimates (representing group differences or covariate effects) across the reduced models, where the weighting is determined by the posterior probability (evidence) of each model. This averaging process ensures that the final parameter estimates and inferences are not conditional on a single ‘best’ model but reflect a consensus across the most plausible explanations supported by the data. Parameters consistently contributing across probable models receive higher weighting, while those unique to less likely models are effectively down-weighted or eliminated.

Finally, inferences within this Bayesian framework are based on the relative evidence supporting the presence versus absence of an effect (e.g., a group difference on a specific connection strength). This is quantified by the posterior probability of the parameter being non-zero. We considered parameters with posterior probabilities exceeding 0.99 as exhibiting strong evidence for an effect.

#### 7.4.1 PEB results

PEB analysis revealed a progressive expansion of synaptic alterations from wake to REM sleep. During wakefulness (supplementary figure 4A), alterations were relatively confined, with the most pronounced difference being decreased NMDA-R-mediated superficial pyramidal self-connections in 22q11.2DS (M = 1.66, SD = 0.99) compared to siblings (M = 2.72, SD = 1.67). AMPA-, GABA-A-, and GABA-B-R-mediated changes were minor during this stage. In N1 (supplementary figure 4B), a stage-dependent reversal was observed, with NMD-RA sp-sp connectivity increased in 22q11.2DS (M = 1.78, SD = 1.18) relative to siblings (M = 1.42, SD = 0.55). N2 sleep (supplementary figure 4C) was marked by a reduction in GABA-A-R-mediated self-inhibition, including si-si (22q11.2DS: M = 1.29, SD = 0.89; siblings: M = 1.52, SD = 0.49), and a tendency toward increased GABA-B-R-mediated inhibition. N3 sleep (supplementary figure 4D) showed a consistent increase in GABA-B-R-mediated inhibition (e.g., dp-dp: M = 1.06, SD = 0.09 vs. M = 0.98, SD = 0.07), while NMDA-R sp-sp connectivity remained lower in 22q11.2DS (M = 1.71, SD = 1.49) than siblings (M = 1.99, SD = 2.22), as in wakefulness. GABA-A and AMPA-R changes were present but less consistent. REM sleep (supplementary figure 4E) exhibited the most extensive alterations. NMDA-R sp-sp connectivity was reduced in 22q11.2DS compared to siblings in REM (M = 2.46, SD = 2.08 vs. M = 2.86, SD = 2.06), consistent with the pattern seen in wake and N3. Exact values for each significant parameter are provided in supplementary table 4.

**Fig S4.**
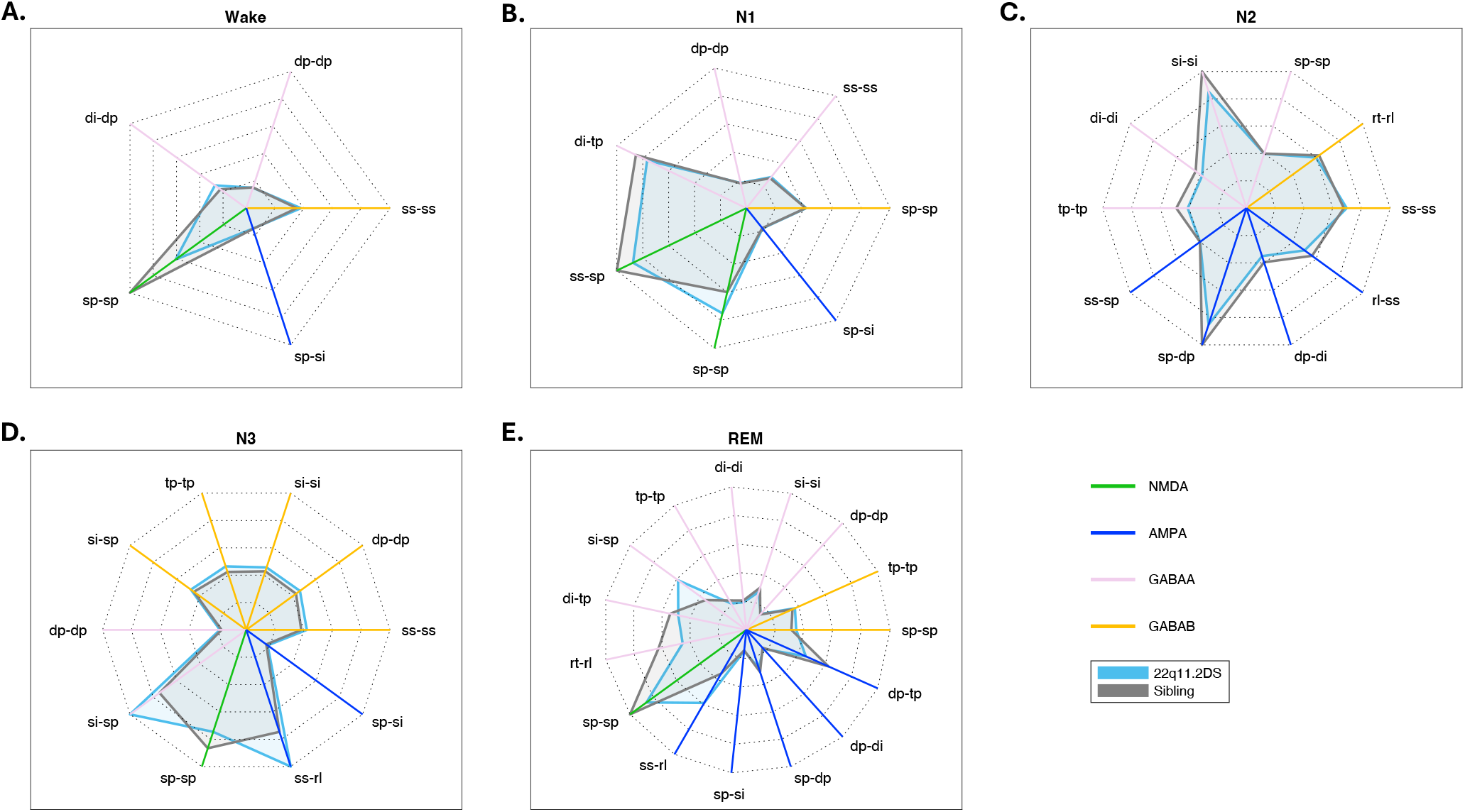
Receptor-specific alterations across sleep-wake states in 22q11.2DS. Each subplot shows the group mean values for parameters with significant group differences (22q11.2DS (blue) vs. sibling controls (grey)) in a given arousal state, as identified by PEB analysis. Each axis radiating from the centre corresponds to a significant parameter, with abbreviations denoting the neuronal populations involved. The colour of each axis indicates the receptor type mediating the connection: NMDA (green), AMPA (blue), GABA-A (pink), and GABA-B (yellow). Only parameters with significant group differences (posterior probability > 0.99) were displayed for each stage.

**Supplementary table 4.** Group means and standard deviations for each significant parameter and sleep stage. The reported means and standard deviations are descriptive summaries of exponentiated parameter estimates within each group and do not reflect the uncertainty of group differences, which are instead inferred using the PEB model. Thus, group distributions may overlap in these descriptive summaries while still indicating strong evidence for a difference in the PEB analysis.

| Sleep Stage | Parameter | 22q11.2DS Mean | 22q11.2DS SD | Sibling Mean | Sibling SD |
| --- | --- | --- | --- | --- | --- |
| Wake | GABA-B ss-ss | 1.036 | 0.317 | 0.957 | 0.133 |
| Wake | GABA-A dp-dp | 0.406 | 0.098 | 0.413 | 0.060 |
| Wake | GABA-A di-dp | 0.735 | 0.435 | 0.607 | 0.194 |
| Wake | NMDA sp-sp | 1.655 | 0.989 | 2.722 | 1.674 |
| Wake | AMPA sp-si | 0.406 | 0.098 | 0.413 | 0.060 |
| N1 | GABA-B sp-sp | 0.984 | 0.136 | 0.977 | 0.127 |
| N1 | GABA-A ss-ss | 0.655 | 0.323 | 0.630 | 0.400 |
| N1 | GABA-A dp-dp | 0.426 | 0.055 | 0.420 | 0.043 |
| N1 | GABA-A di-tp | 1.814 | 0.598 | 2.019 | 0.571 |
| N1 | NMDA ss-sp | 2.073 | 1.110 | 2.368 | 1.658 |
| N1 | NMDA sp-sp | 1.776 | 1.180 | 1.418 | 0.548 |
| N1 | AMPA sp-si | 0.426 | 0.055 | 0.420 | 0.043 |
| N2 | GABA-B ss-ss | 1.058 | 0.192 | 1.028 | 0.174 |
| N2 | GABA-B rt-rl | 0.909 | 0.088 | 0.947 | 0.112 |
| N2 | GABA-A sp-sp | 0.605 | 0.375 | 0.606 | 0.311 |
| N2 | GABA-A si-si | 1.293 | 0.889 | 1.517 | 0.487 |
| N2 | GABA-A di-di | 0.579 | 0.114 | 0.662 | 0.167 |
| N2 | GABA-A tp-tp | 0.620 | 0.187 | 0.740 | 0.295 |
| N2 | AMPA ss-sp | 0.605 | 0.375 | 0.606 | 0.311 |
| N2 | AMPA sp-dp | 1.293 | 0.889 | 1.517 | 0.487 |
| N2 | AMPA dp-di | 0.532 | 0.206 | 0.600 | 0.278 |
| N2 | AMPA rl-ss | 0.758 | 0.260 | 0.855 | 0.350 |
| N3 | GABA-B ss-ss | 0.968 | 0.200 | 0.878 | 0.123 |
| N3 | GABA-B dp-dp | 1.060 | 0.088 | 0.983 | 0.071 |
| N3 | GABA-B si-si | 1.050 | 0.078 | 0.984 | 0.048 |
| N3 | GABA-B tp-tp | 1.068 | 0.133 | 0.973 | 0.087 |
| N3 | GABA-B si-sp | 1.095 | 0.296 | 1.036 | 0.063 |
| N3 | GABA-A dp-dp | 0.422 | 0.071 | 0.395 | 0.097 |
| N3 | GABA-A si-sp | 2.298 | 0.830 | 1.715 | 0.868 |
| N3 | NMDA sp-sp | 1.714 | 1.486 | 1.991 | 2.223 |
| N3 | AMPA ss-rl | 2.298 | 0.830 | 1.715 | 0.868 |
| N3 | AMPA sp-si | 0.422 | 0.071 | 0.395 | 0.097 |
| REM | GABA-B sp-sp | 0.993 | 0.137 | 0.897 | 0.109 |
| REM | GABA-B tp-tp | 1.058 | 0.155 | 1.005 | 0.056 |
| REM | GABA-A dp-dp | 0.427 | 0.125 | 0.412 | 0.096 |
| REM | GABA-A si-si | 0.793 | 0.370 | 0.882 | 0.345 |
| REM | GABA-A di-di | 0.552 | 0.155 | 0.588 | 0.083 |
| REM | GABA-A tp-tp | 0.607 | 0.239 | 0.675 | 0.230 |
| REM | GABA-A si-sp | 1.680 | 0.758 | 1.008 | 0.493 |
| REM | GABA-A di-tp | 1.387 | 0.601 | 1.550 | 0.482 |
| REM | GABA-A rt-rl | 1.297 | 0.888 | 1.789 | 1.079 |
| REM | NMDA sp-sp | 2.459 | 2.078 | 2.863 | 2.061 |
| REM | AMPA ss-rl | 1.680 | 0.758 | 1.008 | 0.493 |
| REM | AMPA sp-si | 0.427 | 0.125 | 0.412 | 0.096 |
| REM | AMPA sp-dp | 0.793 | 0.370 | 0.882 | 0.345 |
| REM | AMPA dp-di | 0.508 | 0.237 | 0.481 | 0.120 |
| REM | AMPA dp-tp | 1.297 | 0.888 | 1.789 | 1.079 |

### 7.5 Randomization test with Monte Carlo approximation

Significance of improvement over the 0% (baseline) adjustment was assessed with a one-sided randomization test with Monte Carlo approximation. For each tested adjustment, we formed two frequency-resolved residual vectors relative to the same mean sibling PSD (baseline vs adjusted group-mean simulation). Under the null hypothesis of within-bin exchangeability, we generated *N* = 1000 independent randomization draws. In each draw, we independently swapped the pair (*d*_adj_(*f*), *d*_base_(*f*)) with probability 1/2, so frequencies were never cross-mixed. For each draw we recomputed MSEs for the relabelled arms and the standardized improvement statistic *T* = (MSE_base_ − MSE_adj_)/MSE_base_, defined as in the main analysis. The one-sided *p*-value was the tail probability under this null, with *N* = 1000 (continuity correction). We report these *p*-values and treat *p* < 0.05 as statistically significant improvement over baseline for this test.

### 7.6 Individual parameter perturbations

To identify the most effective synaptic adjustments for aligning 22q11.2DS neural activity with control patterns, we systematically perturbed individual synaptic parameters and measured their effects on power spectral alignment. For each sleep stage, we analysed the effect sizes and significance of adjustments across different receptor systems and cell-type connections (supplementary figure 5).

**Fig S5.**
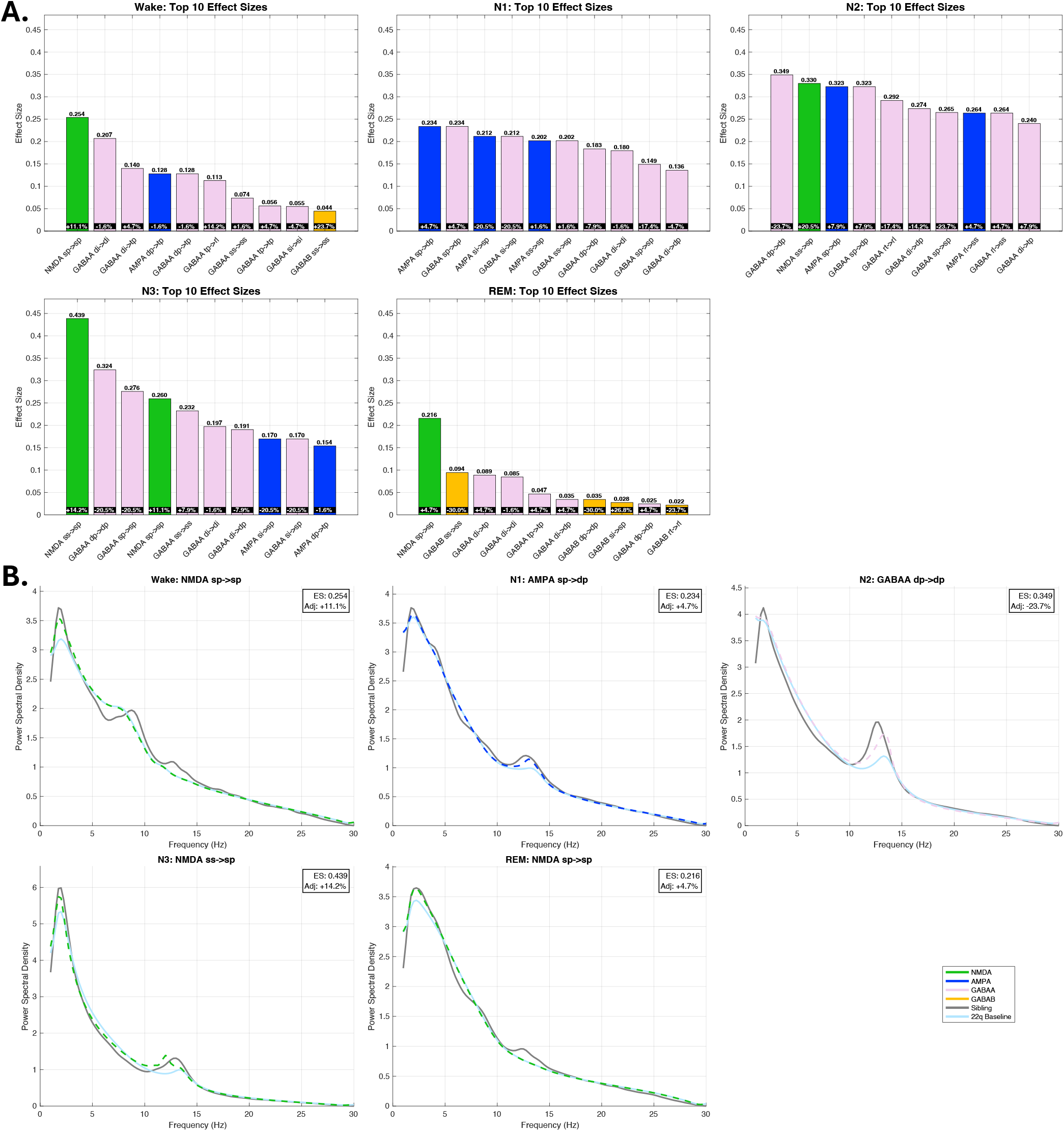
Impact of Receptor-Specific Perturbations on EEG Spectral Power. **A. (Bar Plots):** Each panel represents the single best adjustment (in % with positive values indicating necessary increase and negative values decrease in synaptic gain) for the top 10 receptor-connection pairs that produced a significant improvement in aligning the 22q11.2DS model’s PSD with the sibling control data. For each pair, the optimal adjustment percentage (in the inserted black boxes) and the corresponding maximum effect size are shown. **B. (Power Spectral Density Plots):** Each panel in the bottom row shows the Power Spectral Density (y-axis) across frequencies (x-axis, 0-30 Hz). The light blue line represents the “22q11.2DS Baseline” spectrum. The grey line represents the sibling PSD. The coloured dashed line indicates the receptor type of the perturbation that achieved the overall “best alignment” for that sleep stage. The Effect Size (ES) and Adjustment percentage (Adj) displayed in each PSD plot quantify the effectiveness of this best perturbation in shifting the 22q11.2DS spectrum towards the sibling spectrum.

Notably, NMDA-R sp-sp and NMDA-R sp-ss connections were most consistently among the top parameter adjustments across sleep stages. This finding was robust across both individual connection perturbations and global receptor-wide adjustments (main body section 3.3).

### 7.7 Regression Model Selection for Paramater-Psychiatric/Cognitive Symptom Relationships

Our analysis followed a two-step approach: first using LASSO regression for feature selection, followed by appropriate regression models for the selected features based on their distribution characteristics.

#### 7.7.1 Feature Selection

To manage the high dimensionality of our predictor space relative to our sample size, we first employed LASSO regression^29^ within a nested cross-validation framework to determine the optimal penalty parameter (*λ*). Specifically, we used nested five-fold cross-validation, with LASSO feature selection performed only on training data within each outer fold, and held-out data used to assess out-of-sample predictive performance. A predictor was retained in the final model only if selected in at least 50% of outer folds, ensuring stability across data splits. We acknowledge that final regression coefficients and p-values were estimated by refitting the stage-2 GLM on the full sample using this stable predictor set. Full out-of-sample inference was not feasible given our total sample of N=45, where test folds of approximately nine participants would be insufficient to reliably fit negative binomial or logistic models. Results from the stage-2 models should therefore be interpreted as exploratory.

#### 7.7.2 Regression Models for Selected Features

Following LASSO feature selection, we analysed the relationships between selected features and outcome measures using appropriate regression models based on their distribution characteristics:

For psychiatric measures, count data (ADHD, anxiety, ASD symptoms, and sleep disturbances) were first tested for overdis-persion. When the variance significantly exceeded the mean (p < 0.05), indicating overdispersion, we used negative binomial regression instead of Poisson regression. For all our count measures, overdispersion was present, leading to the use of negative binomial models. Psychotic experiences were analysed using logistic regression as this was a binary outcome measure.

For cognitive measures, Working Memory and SOC Problems Solved were analysed using linear regression after confirming normality using Shapiro-Wilk tests^90^ (p > 0.05) and Q-Q plots. Processing Speed and SOC Initial/Subsequent Time measures required log transformation, before linear regression analysis. RVP A’, being bounded between 0 and 1 and showing no significant deviation from normality (Shapiro-Wilk test, p > 0.05), was analysed using linear regression. Perseverative and Nonperseverative Errors showed significant overdispersion in initial Poisson models and were therefore analysed using negative binomial regression (supplementary figure 6).

**Fig S6.**
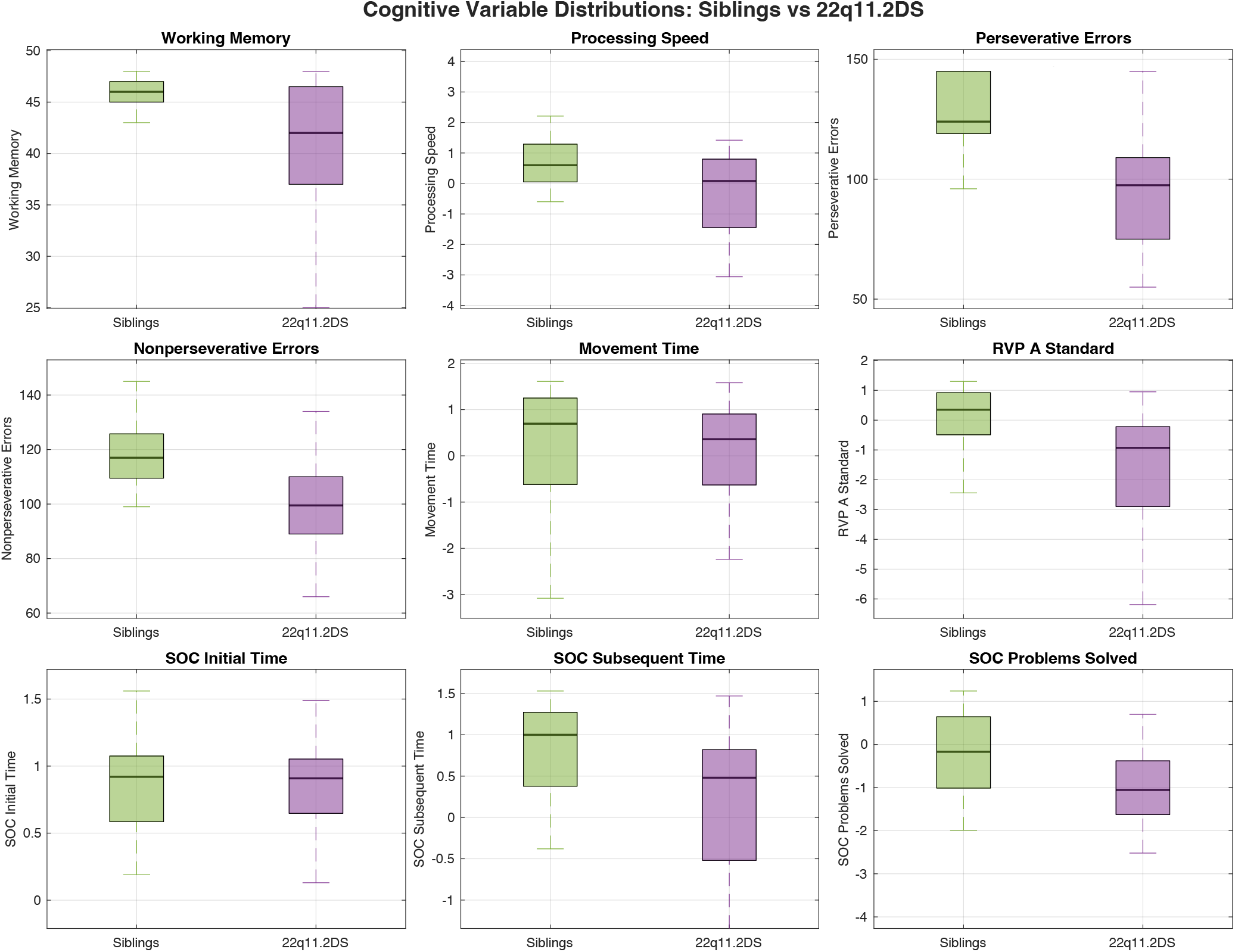
Cognitive Variable Distributions in 22q11.2DS and Siblings. Box plots showing the distribution of cognitive measures between 22q11.2DS carriers (purple) and sibling controls (green). Measures include working memory performance, processing speed, perseverative and nonperseverative errors (executive function), movement time, sustained attention (RVP A’ Standard), and planning ability (SOC Initial Time, Subsequent Time, and Problems Solved). The boxes show the interquartile range with the median line, whiskers extend to 1.5 times the interquartile range.

## Notes

### Competing Interest Statement

The authors have declared no competing interest.

### Author Declarations

This study did not generate any new data. All analyses were conducted on previously published data from the IMAGINE-ID/ECHO study, available as part of the original publication by Donnelly et al. (2022). Source Data files are included with that publication. Nicholas A Donnelly Ullrich Bartsch Hayley A Moulding Christopher Eaton Hugh Marston Jessica H Hall Jeremy Hall Michael J Owen Marianne BM van den Bree Matt W Jones (2022) Sleep EEG in young people with 22q11.2 deletion syndrome: A cross-sectional study of slow-waves, spindles and correlations with memory and neurodevelopmental symptoms eLife 11:e75482. https://doi.org/10.7554/eLife.75482

### Summary of Updates

The ORCID of Jeremy Hall which accidentally was linked to another Jeremy Hall employed by Cardiff University.

